# Health aspects of vegan diets among children and adolescents: a systematic review and meta-analyses

**DOI:** 10.1101/2023.05.07.23289579

**Authors:** Alina Koller, Sabine Rohrmann, Maria Wakolbinger, Jan Gojda, Eliska Selinger, Monika Cahova, Martin Svetnicka, Sandra Haider, Sabrina Schlesinger, Tilman Kuhn, Jeffrey Keller

## Abstract

Health effects of vegan diets among children are a controversial public health topic. Thus, we evaluated a broad range of health outcomes among vegan children and adolescents aged 0 to 18 years by a systematic review and meta-analyses. 17 studies met the inclusion criteria (16 cross-sectional studies, one randomized trial). Meta-analyzes showed lower intakes of protein (%E MD[95%CI]: −3.54 [−5.08, −2.00]) and higher intakes of fiber (g/1000kcal MD[95%CI]: 8.01 [6.96, 9.06]) in vegans compared to omnivores. Non-meta-analyzable studies showed lower calorie, vitamin B12 (when not using supplements) and selenium intakes, as well as lower blood levels of ferritin and vitamin B12. By contrast, vegans had significantly higher intakes of folate, vitamin C and iron. Levels of several cardiometabolic biomarkers (cholesterol, LDL) and indicators of bone health (vitamin D and calcium intake, vitamin D blood levels, bone mineral density) were lower in vegans. Risk of Bias was rated as high or very high in seven out of 17 primary studies. The Certainty of the Evidence was low or very low in all meta-analyses. Vegan children and adolescents following a vegan diet may have a beneficial cardiovascular risk profile but may be at risk for impaired bone health.

## Introduction

Plant-based diets are gaining strong interest for individual and planetary health reasons. Some of these diets, i.e. pescetarian (characterized by an exclusion of meat, but a consumption of fish as well as eggs and dairy), lacto-ovo-vegetarian (no meat and fish, but eggs and dairy), or lacto-vegetarians (no meat, fish, and eggs, but dairy) diets, still contain animal products, while a vegan diet excludes all animal products (1). In recent years, veganism has become particularly popular (2,3). This may be explained by the medial presence of issues such as global warming, industrial livestock farming, animal rights and the potential negative health effects of animal foods (4). A market research report from 2017 indicated that 6% of US consumers claimed to be vegan (5). According to a survey by *Euromonitor International*, 3.4% of Europeans reported following a vegan diet in 2021 (6).

As veganism becomes more popular, more and more parents are considering a vegan diet for their children. Childhood is a vulnerable life stage in terms of nutritional deficiencies, as children are still growing (7) and developing (8,9). Therefore, it is essential to assess the risk and possible benefits of a vegan diet during childhood and adolescence. However, most previous studies on health outcomes among vegans were conducted among adults. A systematic review from 2018 showed lower intakes of total energy, protein and fat but a higher intake of carbohydrates and polyunsaturated fatty acids among vegan adults (10).A recent published umbrella review on various health outcomes in adults suggested that a vegan diet may reduce cardiometabolic risks but impact bone health (11). A literature review on studies among children published in 2021 indicated a lower calcium, vitamin D and cobalamin intake among vegan children (aged 4 months to 10 years) (12). Generally, primary studies concerning a vegan diet among children and adolescents are scarce and, up to now, the findings have not been summarized systematically yet.

National dietary guidelines include different recommendations concerning a vegan diet among children and adolescents. For example, the Swiss Nutrition Society does not recommend a vegan diet in childhood. If parents still decide to provide a vegan diet to their children, supervision by a pediatrician is recommended (13). The German Nutrition Society expressed concerns about nutrient and vitamin deficiencies among vegan children in a position statement from 2016 (14). The British Nutrition Foundation and the Czech pediatric society also expressed concerns about a vegan diet for children and newborns (15,16), whereas the Canadian Paediatric Society declares that a well-balanced vegan diet is suitable for children and adolescents (17). The US Academy of Nutrition and Dietetics (18), the Portuguese National Programme for the Promotion of a Healthy Diet (19) and the Australian National Health and Medical Research Council (20) all state that a well-planed vegan diet is healthful and adequate for all stages of life, including childhood. These differences in recommendations may be explained by different interpretations of the limited number of studies concerning a vegan diet among children.

Given the growing interest in veganism and contradictory national guidelines, we carried out a systematic review of human studies on health outcomes among vegan children and adolescents.

## Methods

The systematic literature research was performed following the preferred reporting items for systematic reviews and meta-analyses (PRISMA guidelines) (21). The study protocol was registered at PROSPERO (CRD42022314594).

### Literature search

The systematic literature search was performed in PubMed and Embase up to April 2023. The detailed search terms are presented in *supplementary Table S1 and S2*. No restrictions or filters where applied. The literature research and screening of the articles were performed by two independent researchers (AK, JK) to ensure replicability. In addition to the systematic literature research, a “hand search” was conducted, using the literature list of other systematic reviews concerning vegan diets. All studies were uploaded to Cadima, which was used for the study selection (22). Cross-sectional studies and prospective studies including randomized controlled trials investigating the association between a vegan diet defined as a plant-based diet avoiding any product of animal origin (except human breastmilk) for at least one year/ since birth and any health outcomes among children and adolescents 0 to 18 years of age were eligible. The comparison group were omnivorous children and adolescents. Our PICO statement is presented in *supplementary Table S3*.

## Data extraction and assessment of risk of bias

### Data extraction and analysis

One researcher (AK) extracted the data from the primary studies and another researcher (JK) double-checked it for accuracy. For each primary study, the following data were extracted: Name of the first author, publication year, country, study design, definition of the exposure, definition of the outcome, total number of participants for both comparison vs. intervention group separately, age range, means and standard deviation. If studies presented the results as quantiles instead, means and standard deviation were calculated in R (version 4.2.2) using the “estmeans” package(v1.0.0; function bc.mean.sd) (23). If a particular outcome was meta-analyzable, the data was analyzed with RevMan5. If multiple studies reported on the same outcome but the age groups were too heterogeneous, we visualized the between group effect sizes in a heat map (R package ggplot2; version 3.4.0) (24). Effect sizes and corresponding p-values were calculated using differences between group means and SDs of both groups where present (Welch’s standard error).

### Assessment of the risk of bias

For the assessment of the risk of bias, ROBINS-E (tool to assess risk of bias in non-randomized studies of exposures) (25) was used for observational studies and the RoB-2.0 tool for RCTs (26). ROBINS-E and RoB-2.0 assessments were performed by two researchers (AK, JK) independently for each study. Any discrepancies were discussed with a third researcher (TK) and resolved by consensus. For further information, see *supplementary Table S7* and *supplementary information about ROBINS-E/RoB 2.0*.

## Evaluation of quality of evidence

GRADE was used for evaluating the quality of evidence of the outcomes that were meta-analyzed in this review (27). GRADE is a scoring system (high, moderate, low, very low) that rates five factors (*study limitations/ risk of bias, inconsistency, indirectness, imprecision, publication bias)* for downgrading and two factors (*large effect and dose response effect)* for upgrading. For more detailed information see *supplementary Table S8* and *supplementary information about GRADE*.

## Results

### Literature Search

A total of 2346 publications were identified, of which 406 were duplicates. Of the 1940 publications that were screened for title and abstracts, 62 were included for full text screening. 45 studies were excluded for pre-specified reasons i.e., as not fitting in the PICO scheme, duplicates, or due to unavailable full texts or supplements. Overall, 17 reports were included in this systematic review. The flow chart of the study selection progress is shown in *supplementary Figure S1* and the list of excluded studies in *supplementary Table S4.* Study characteristics (year, country, participants, adjusted confounders) are presented in *supplementary Table S5*. An overview of all extracted results (including recalculated confidence intervals, means and SD’s) are presented in *supplementary Table S6*.

Overall, nine meta-analyses with the following outcomes were conducted: intakes of relative carbohydrate, fiber, protein, fat, polyunsaturated fatty acid, monounsaturated fatty acid, and saturated fatty acid as well as birth weight and BMI percentiles. Although more than nine outcomes were examined by more than two studies, many of them were not suitable for meta-analyses because the age groups of the study participants were too heterogeneous and results correlated with age. Differences in statistical effect sizes across ages are visualized for different endpoints in *Supplementary Figure S2*. Thus, heatmaps of the corresponding outcomes were created, in which effect sizes from individual studies are shown across study populations with different age ranges (Figure 1). The directions of the associations between vegan diet and health outcomes across the included studies are presented in Table 1-4.

**Figure 1.**
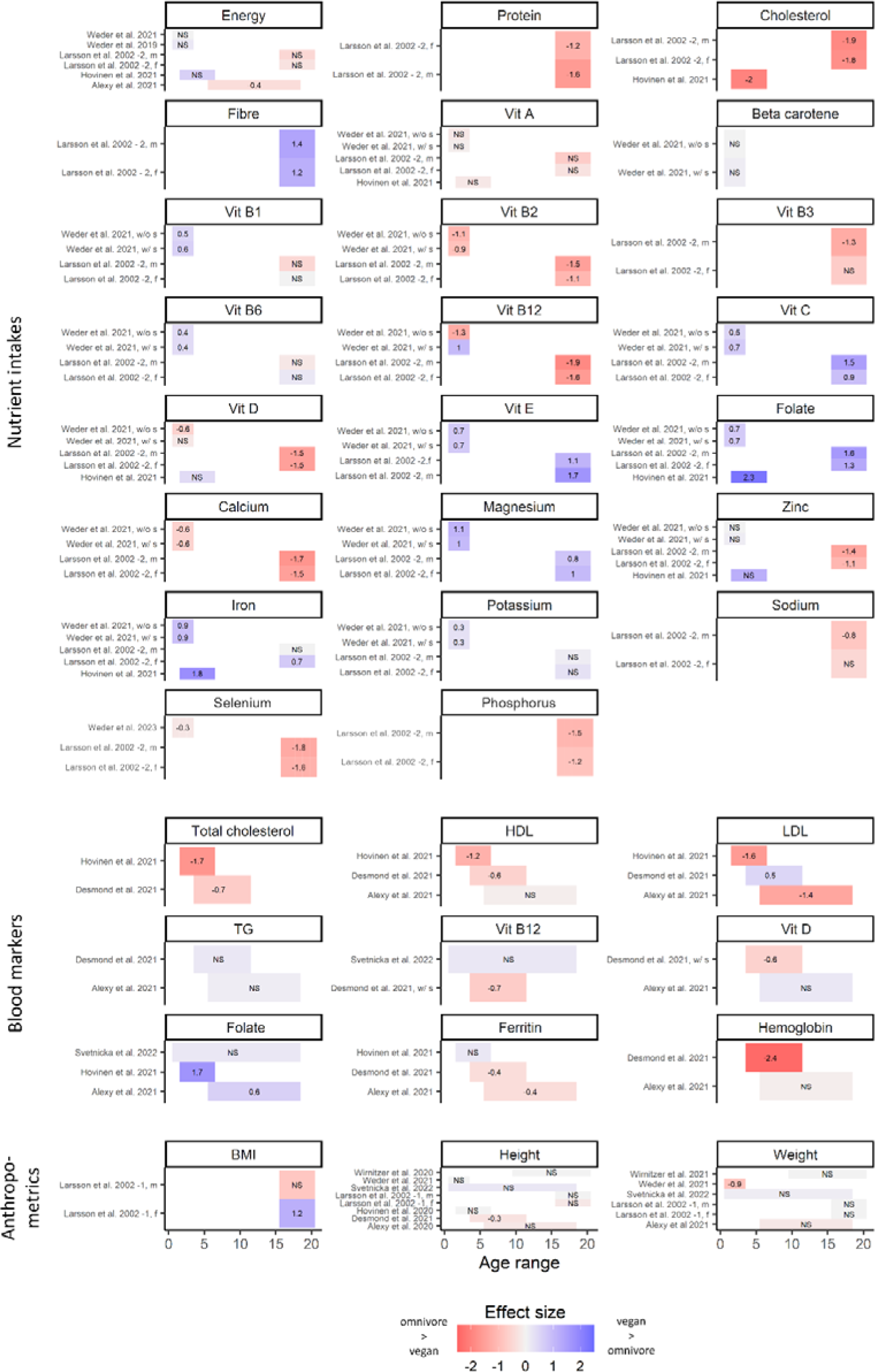
heat map summarizing effect sizes of different outcomes; Vit, Vitamin; LDL, low-density-lipoprotein; HDL, high-density-lipoprotein; TG; triglycerides; BMI, body mass index; f, female; m, male; w/s, with supplement; w/o s, without supplement; NS, not significant

**Table 1.**
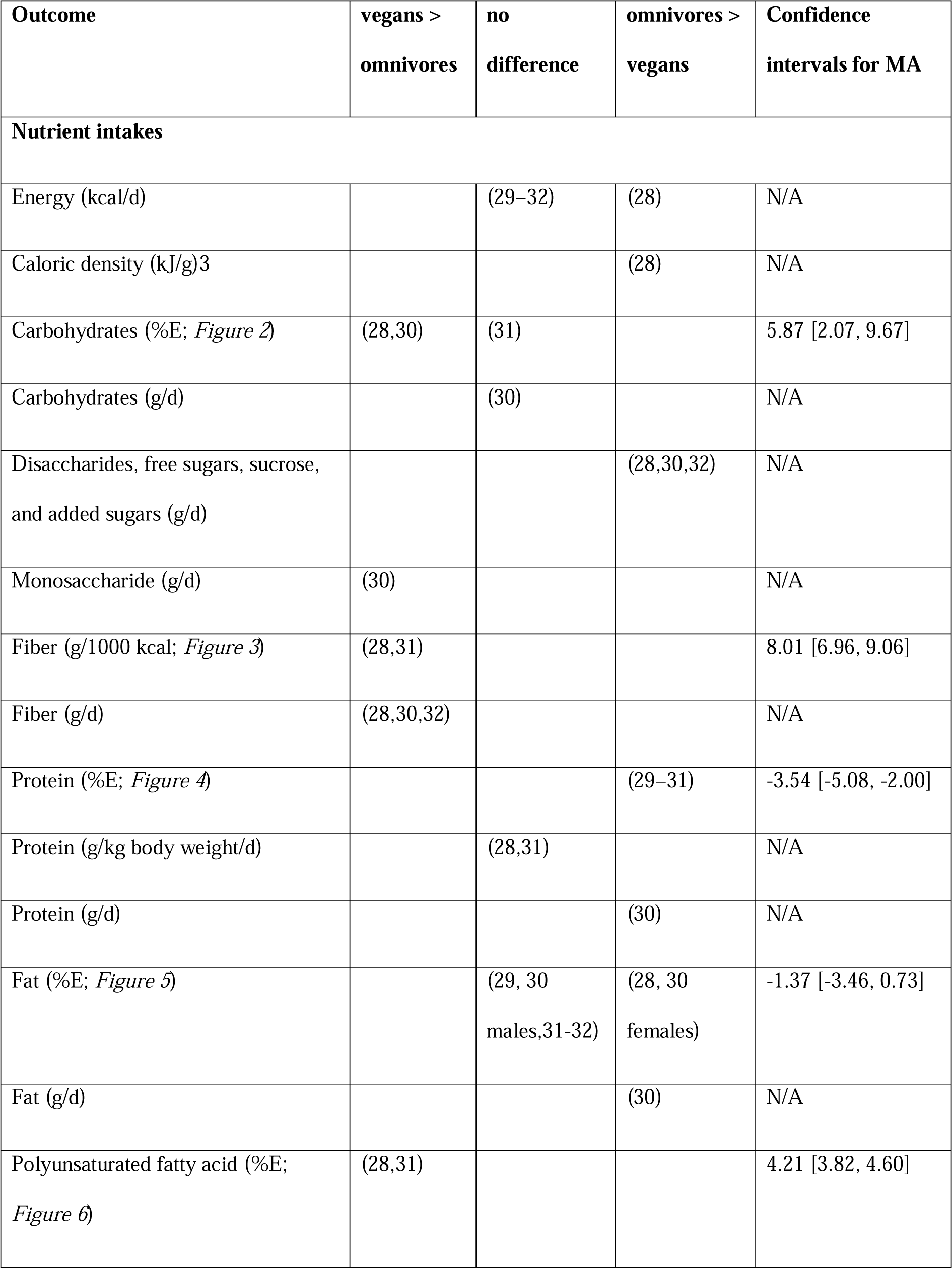
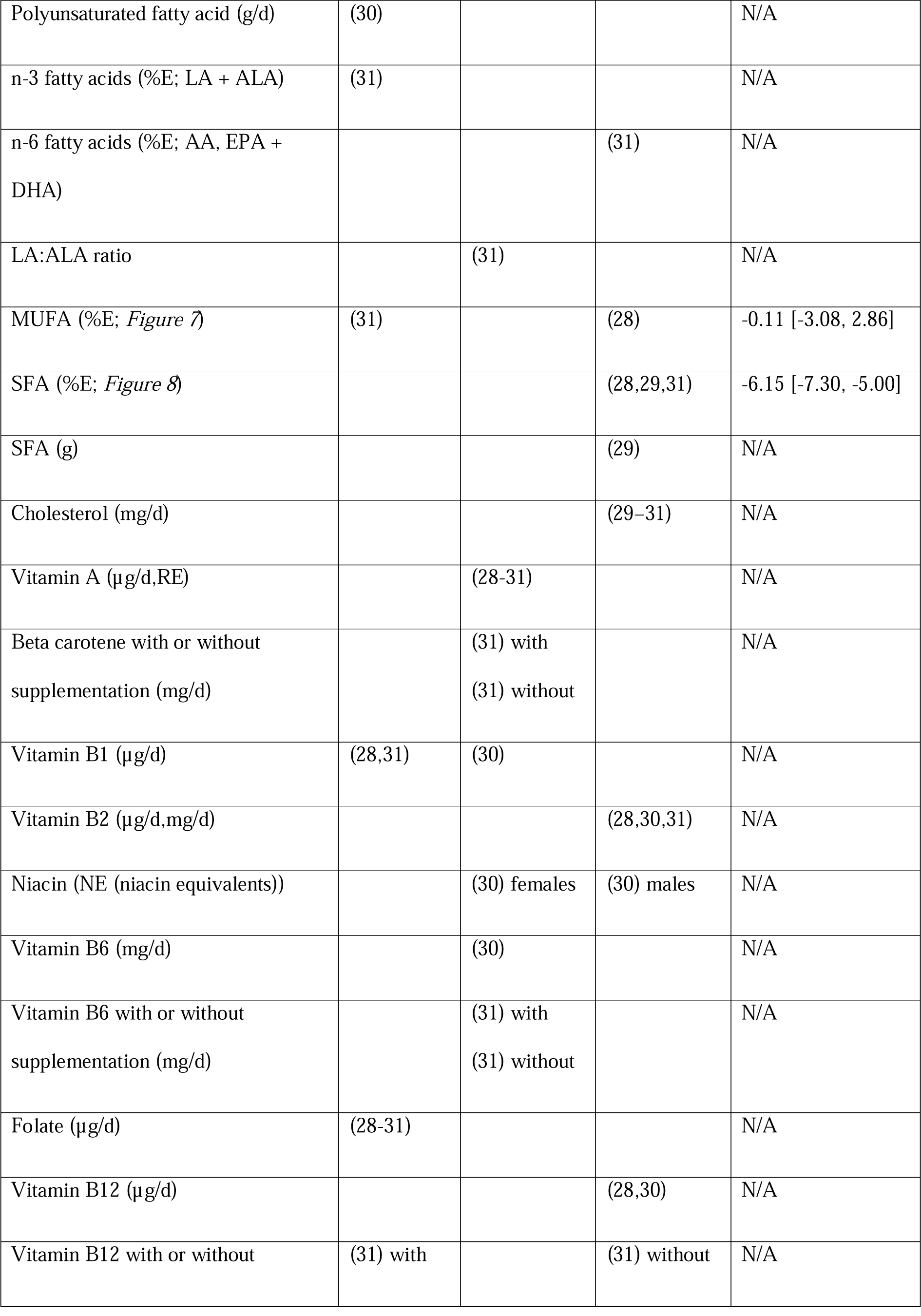
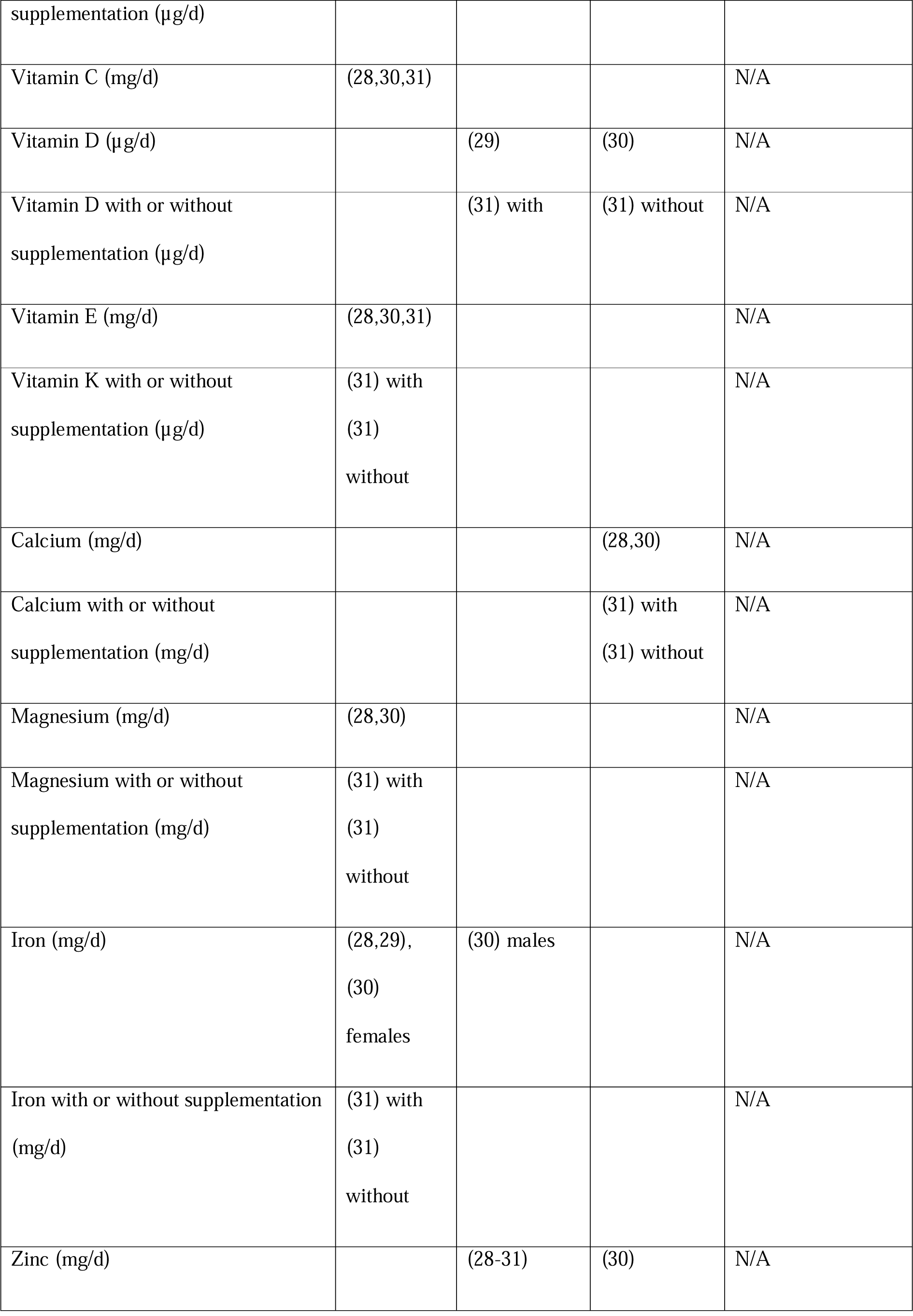
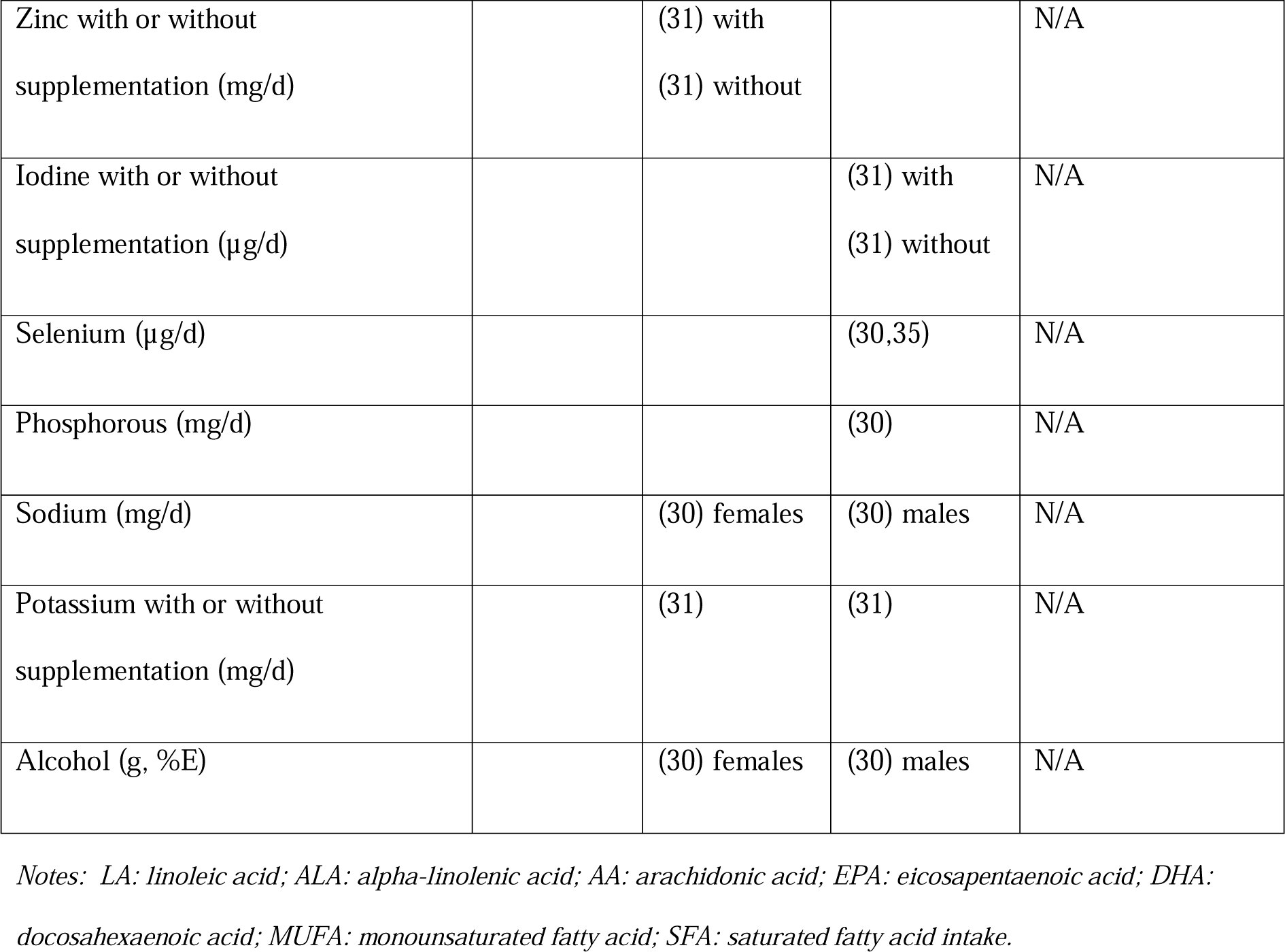
Overview of nutrient intake differences in vegan compared to omnivorous children.

Results of the meta-analyses are shown in **Figures 2-10**. The majority of the meta-analyses was on relative outcomes (nutrients in % Energy intakes, g/1000kcal) and anthropometric parameters (BMI and birth weight) in percentiles as these units are comparable across the different age groups (detailed information about age ranges in *supplementary Table S3*).

**Figure 2.**
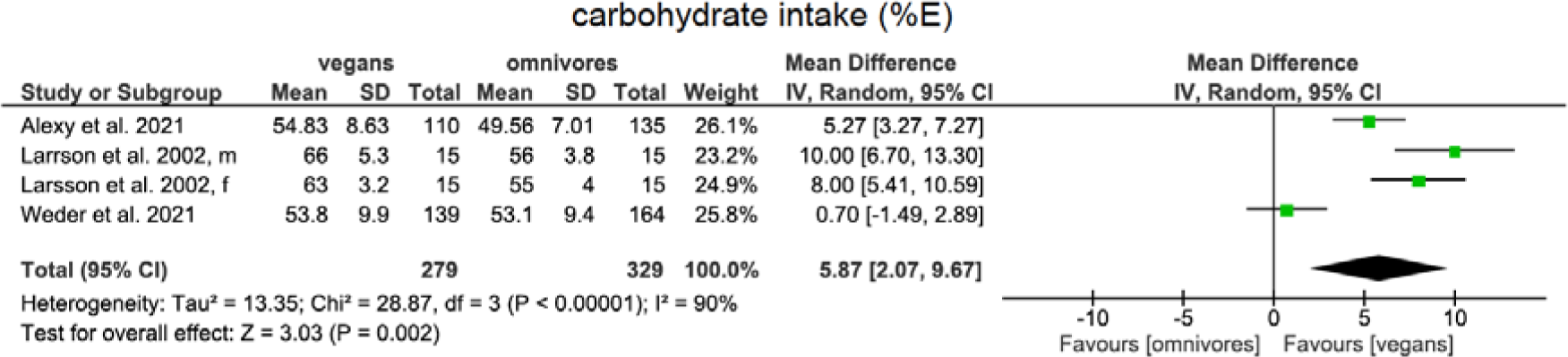
Mean differences with 95CIs using a random effects model for a vegan diet compared to an omnivore diet regarding carbohydrate intake in %Energy among children

#### Macronutrient intake and status

An overview of all outcomes concerning nutrient, vitamin and mineral intake and status are presented in Tables 1 and 2.

**Table 2.** Overview of differences in nutrient blood concentrations in vegan compared to omnivorous children.

| Outcome | vegans ><br>omnivores | no<br>difference | omnivores ><br>vegans | Confidence<br>intervals for MA |
| --- | --- | --- | --- | --- |
| **Nutrient blood concentrations** |  |  |  |  |
| ALA (log2FC) | (29) |  |  | N/A |
| DHA (log2FC) |  |  | (29) | N/A |
| 20:3n-6, 22:4n-6 (in erythrocytes) | (49) |  |  | N/A |
| DHA (in erythrocytes) |  |  | (49) | N/A |
| Cholesterol (mmol/l, mg/dl) |  |  | (29,33) | N/A |
| Vitamin A (arbitrary units) |  | (29) |  | N/A |
| Vitamin B2 (µg/l) |  |  | (28) | N/A |
| Folate (µg/l, nmol/l) | (28,29) | (34) |  | N/A |
| Vitamin B12 (µg/l, pmol/l) | (34) |  |  | N/A |
| Vitamin B12 with or without<br>supplementation (pmol/l) |  | (33) with | (33) without | N/A |
| Homocysteine with or without<br>supplementation (%) | (33) with<br>(33)<br>without |  |  | N/A |
| Holotranscobalamin (pmol/l) | (28) |  |  | N/A |
| Vitamin D (ng/ml) |  | (28) |  | N/A |
| Vitamin D with or without supplementation (nmol/l) |  | (33) with | (33) without | N/A |
| Ferritin (% , µg/l) |  | (29) | (28,33) | N/A |
| RBC (M/µl) |  |  | (33) | N/A |
| Hemoglobin (g/dl) |  |  | (31) | N/A |
| Platelets (mg/dl) |  |  | (28) | N/A |
| MCV (fl) |  | (34) |  | N/A |
| MCV with or without supplementation (fl) | (33) with<br>(33)<br>without |  |  | N/A |
| Zinc (µmol/l) |  | (29) |  | N/A |
| Iodine /creatinine urinary (µg/l per mmol/l) |  | (29) |  | N/A |

*Notes: LA: linoleic acid; ALA: alpha-linolenic acid; AA: arachidonic acid; EPA: eicosapentaenoic acid; DHA: docosahexaenoic acid; MCFA: medium-chain fatty acid; MUFA: monounsaturated fatty acid; SFA: saturated fatty acid intake; RBC: red blood cell count; MCV: mean corpuscular volume;*

Energy intake was lower in vegan children compared to omnivores in one study (28). Four studies (29–32) found no difference in caloric intake per day. One study (28) found a lower caloric density (kcal/g food) in vegan compared with omnivorous children.

Relative carbohydrate intake (%E) was found to be higher among vegan children in the meta-analysis including three primary studies (mean difference, MD[95%CI]:5.87 [2.07, 9.67], very low Certainty of Evidence (CoE)) (see Figure 2) (28,30,31). A further study among children (30) showed no difference in absolute carbohydrate intake compared to omnivore children.

Fiber intake (g/1000kcal) was assessed in a meta-analysis of two studies (28,31), which showed a higher intake among vegan children (MD[95%CI]: 8.01 [6.96, 9.06], very low CoE) (see Figure 3). Three other studies that could not be meta-analyzed (28,30,31) showed higher absolute intakes of fiber (g/d) in vegans compared with omnivores.

**Figure 3.**
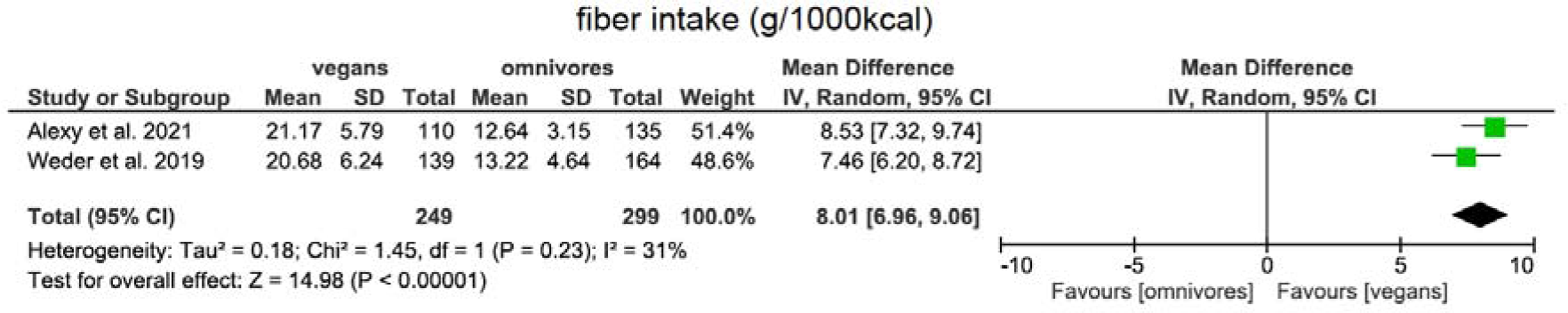
Mean differences with 95CIs using a random effects model for a vegan diet compared to an omnivore diet regarding fiber intake in g/1000kcal among children.

Protein intake (%E) was assessed in a meta-analysis of three studies (29–31) and revealed a lower intake among vegan children (MD[95%CI]: −3.54 [−5.08, −2.00], very low CoE) (see Figure 4). With respect to protein intake in proportion to body weight (g/kg BW/d), there was no difference in two studies whereas absolute protein intake (g/d) was lower among vegans in one study (30).

**Figure 4.**
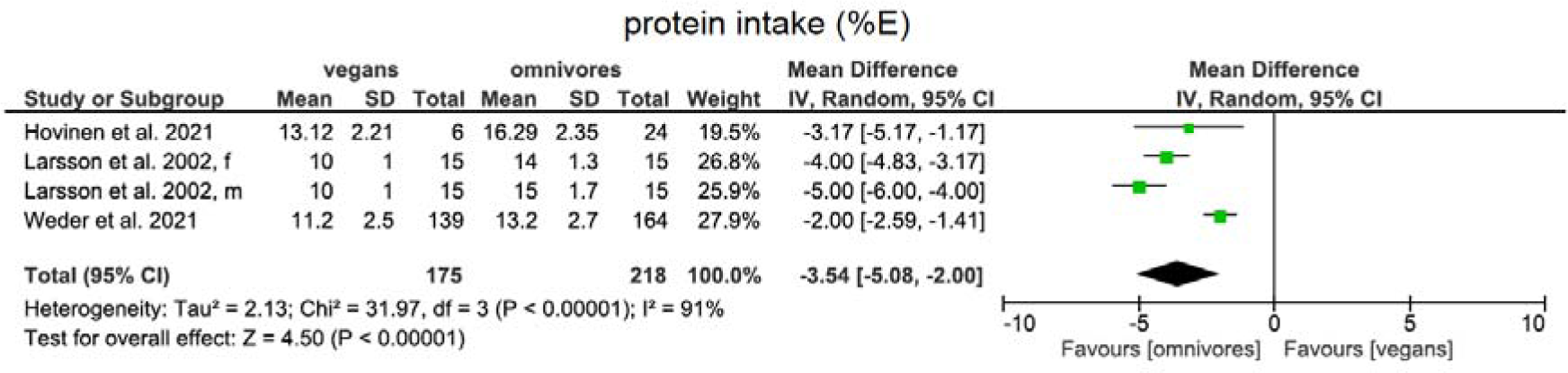
Mean differences with 95CIs using a random effects model for a vegan diet compared to an omnivore diet regarding protein intake in % Energy among children.

Fat intake (%E) was analyzed in a meta-analysis of five studies and showed no difference between the two groups (MD[95%CI]: −1.37 [−3.46, 0.73], very low CoE) (28–32) (see Figure 5). One of the meta-analyzed studies, additionally evaluated absolute fat intake, which was lower in vegans compared to omnivores (30).

**Figure 5.**
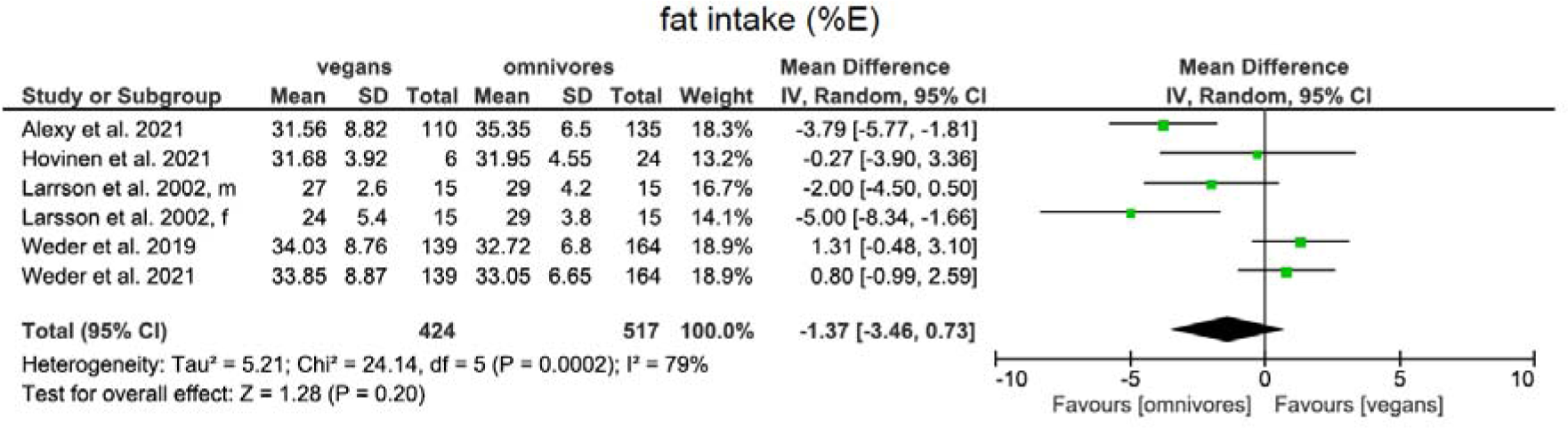
Mean differences with 95CIs using a random effects model for a vegan diet compared to an omnivore diet regarding fat intake in %Energy among children.

Polyunsaturated fatty acid intake (%E) was assessed in a meta-analysis of two studies (28–32) and showed a higher intake among vegan children (MD[95%CI]: 4.21 [3.82, 4.60], very low CoE) (see Figure 6). A study by Larsson et al., which was not included in the meta-analysis, observed higher intakes in vegan children in both sexes (30).

**Figure 6.**
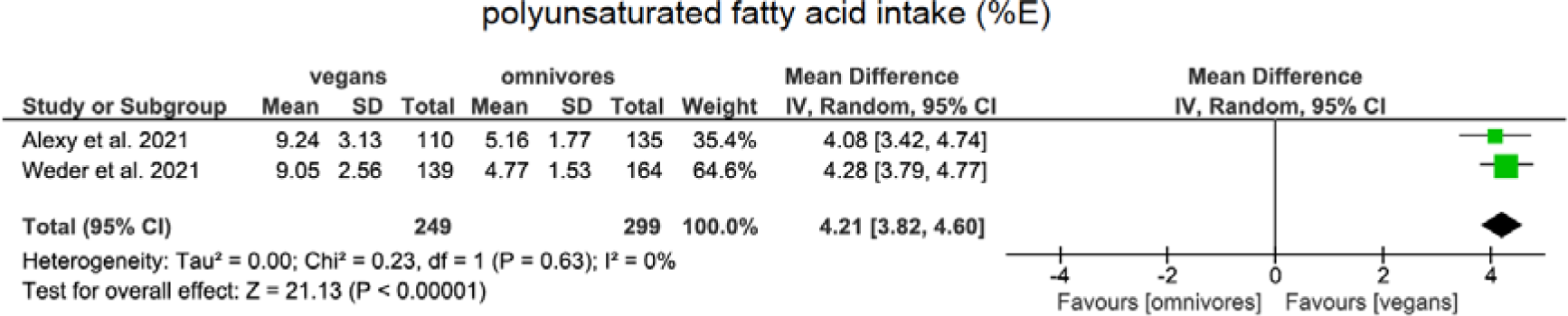
Mean differences with 95CIs using a random effects model for a vegan diet compared to an omnivore diet regarding polyunsaturated fatty acid intake in %Energy among children.

Monounsaturated fatty acid (MUFA) intake (%E) was meta-analysed based on two studies (28–31), which showed no clear difference (MD[95%CI]: −0.11 [−3.08, 2.86], very low CoE) (see Figure 7).

**Figure 7.**
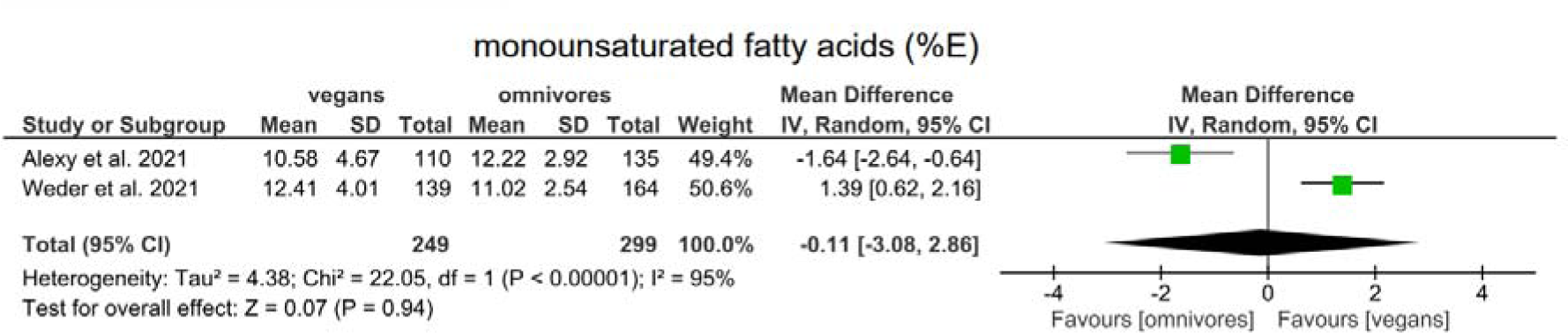
Mean differences with 95CIs using a random effects model for a vegan diet compared to an omnivore diet regarding monounsaturated fatty acid intake in %Energy among children.

Saturated fatty acid intake (SFA) (%E) was assessed in a meta-analysis of three studies (28,29,31) and showed a lower intake among vegan children (MD[95%CI]: −6.15 [−7.30, - 5.00], very low CoE) compared with omnivore children (see Figure 8). Another study (29) showed a lower absolute SFA (g) intake in vegans compared to omnivores.

**Figure 8.**
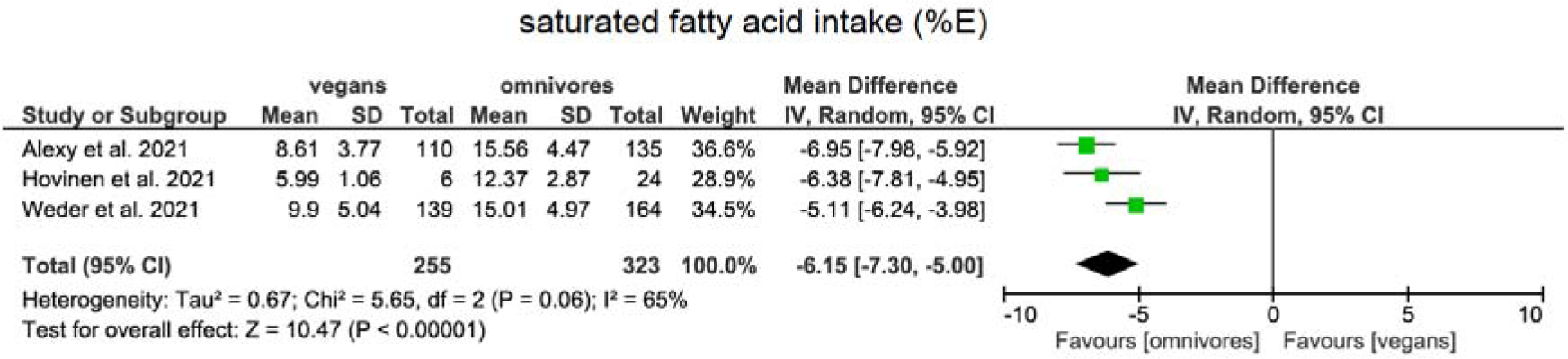
Mean differences with 95CIs using a random effects model for a vegan diet compared to an omnivore diet regarding saturated fatty acid intake in % Energy among children.

Cholesterol intake was lower among vegans in all three included studies (29–31). Two studies (29,33) examined blood cholesterol levels, which were lower in vegans in both studies.

#### Single study findings concerning nutrient intake and status

Disaccharides, free sugars, sucrose, added sugars intake (30,32,33), EPA, AA, DHA intake (33) DHA blood levels (31) cholestanol, campesterol, sitosterol and avenasterol (29) and alcohol intake (only in males) (30) were lower in vegans compared to omnivores. LA:ALA (33) ALA levels (31), squalene, desmosterol, cholestenol and lathosterol (29) levels showed no difference between vegan and omnivore children. Monosaccharide intake (30), LA and ALA intake (31) were higher in vegans compared to omnivores.

#### Vitamin and mineral intake and status

Both blood concentration and intakes of vitamin A and beta carotene were not different between the two groups (28-31), with the exception of one study (29) that found a lower vitamin A concentration in vegans compared with omnivores.

Vitamin B1/thiamin intake was examined in two studies (28,31) and was higher among vegans. Another study (30) found no difference. Vitamin B2/riboflavin intake was lower in vegans in three studies (28,30,31). Blood concentration was determined in only one study (28) and was lower among vegans as well. Vitamin B6 intake was significantly higher among vegans in one study (30). A second study (31) showed no significant difference. Folate intake was higher in vegans in all four studies included (28–31). Two of three studies that determined blood folate levels showed higher concentrations in vegans (28,29).

Vitamin B12 intake was lower in vegan children in two studies (28,30). One study distinguished between vitamin B12 intake with and without supplementation (31) and showed lower intakes in vegan children without supplementation than in the omnivore comparison group. With supplementation, intakes were higher in vegans than in omnivores. Two studies examined blood concentrations of vitamin B12 (33,34). The first study found a higher level of vitamin B12 among vegans compared to omnivores. The second study distinguished between blood levels with and without supplementation (33). Without supplementation vegans had lower blood levels than omnivores, with supplementation there was no difference.

Vitamin C intake was higher in vegans in all three included studies (28,30,31).

Vitamin D intake was assessed in three studies. One study found no difference (29). Weder et al. examined vitamin D intake both with and without supplementation (31). Dietary vitamin D intake with supplementation did not differ between vegan and omnivores. The intake without supplementation was lower in the vegan children compared to omnivores. Another study (30) observed a lower vitamin D intake in both sexes in vegans than in omnivores. Desmond et al. examined blood vitamin D levels both with and without supplementation. Without supplementation, the blood level of the vegan group was lower, with supplementation there was no difference (33). Another study showed no difference in blood levels of vitamin D between vegans and omnivores (28).

Calcium intake was lower and magnesium intake was higher in vegans than in omnivores in all three identified studies (28,30,31).

In three studies, iron intake was higher in vegans than in omnivores (28,29,31). Another study (30) found a higher intake in vegans but only in females. Two of three studies (28,33) that examined blood ferritin levels found lower levels in vegans.

Zinc intake was lower in both sexes in one study (30) in the vegan children. Three studies each showed no difference in zinc intake (28-31) and blood levels of zinc (28,29).

Iodine intake was lower among vegans with and without supplementation in one study (31). The study by Hovinen et al. examined urinary iodine levels and showed no difference (29).

Selenium intake was significantly lower among vegans than omnivores in two studies (30,35).

#### Single study findings concerning vitamin and mineral intake and status

Niacin (only in males), phosphorus, sodium (only in males) intakes were lower among vegans compared to omnivores (30).

#### Anthropometric parameters

All results concerning anthropometric parameters are presented in Table 3.

**Table 3.** Overview of anthropometric differences in vegan compared to omnivorous children.

| Outcome | vegans ><br>omnivores | no<br>difference | omnivores ><br>vegans | Confidence<br>intervals for MA |
| --- | --- | --- | --- | --- |
| **Anthropometric parameters** |  |  |  |  |
| Height (percentile or z score) |  | (28,31) | (33) | N/A |
| Height (cm) |  | (28,29,39) | (31,36) | N/A |
| Head circumference (cm/percentile)<br>birth, 6,12 months |  | (37) |  | N/A |
| Skinfolds (subscapular, biceps),<br>waist girth |  | (33) |  | N/A |
| Skinfolds (suprailiac,triceps) tight-,<br>hip girth |  |  | (33) | N/A |
| Body weight (kg) | (39)<br>females | (28,29),(39)<br>males | (31,36) | N/A |
| Weight for age & weight for height<br>score |  | (31) |  | N/A |
| Birth weight (kg; *Figure 9*) | (38) | (37) | (33) | 0.08 [-0.21, 0.38] |
| BMI percentiles (*Figure 10*) |  | (34,36) |  | 1.60 [-0.86, 4.06] |
| BMI (kg/m<sup>2</sup>) | (39) males | (39) males |  | N/A |
| BMI z-score |  | (28,29) | (33) | N/A |

Notes: BMI: body mass index.

Two studies (28,31) compared height by percentile or z score, showing no difference between vegan and omnivore children. One study (33) observed lower values for vegans. In two of five studies that compared absolute height (31,36), the vegan children were shorter, but there were no differences in the other three studies.

Body weight was examined in five studies. Two studies (28,29) showed no difference in body weight between the vegan and non-vegan groups. In one study (30), body weight was higher in vegan girls compared to omnivore girls, whereas there was no difference for boys. Two studies found a lower body weight in vegan children compared to omnivore children (31,36).

Birth weight was assessed in a meta-analysis including three studies (33,37,38) and showed no statistically significant difference (MD[95%CI]: 0.08 [−0.21, 0.38], p=0.58, very low CoE) (see Figure 9).

**Figure 9.**
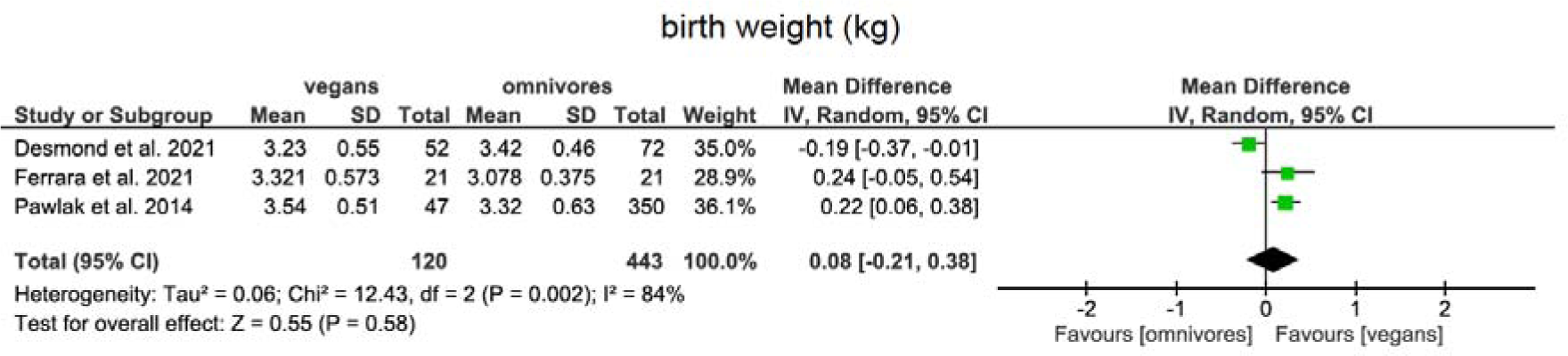
Mean differences with 95CIs using a random effects model for a vegan diet compared to an omnivore diet regarding birth weight in kg.

BMI percentiles were presented in a meta-analysis including two studies (34,36) and showed no difference between the two groups (MD[95%CI]: 1.60 [−0.86, 4.06], low CoE) (see Figure 10). One study (30) showed higher BMI in vegan girls but with no difference in boys.

**Figure 10.**
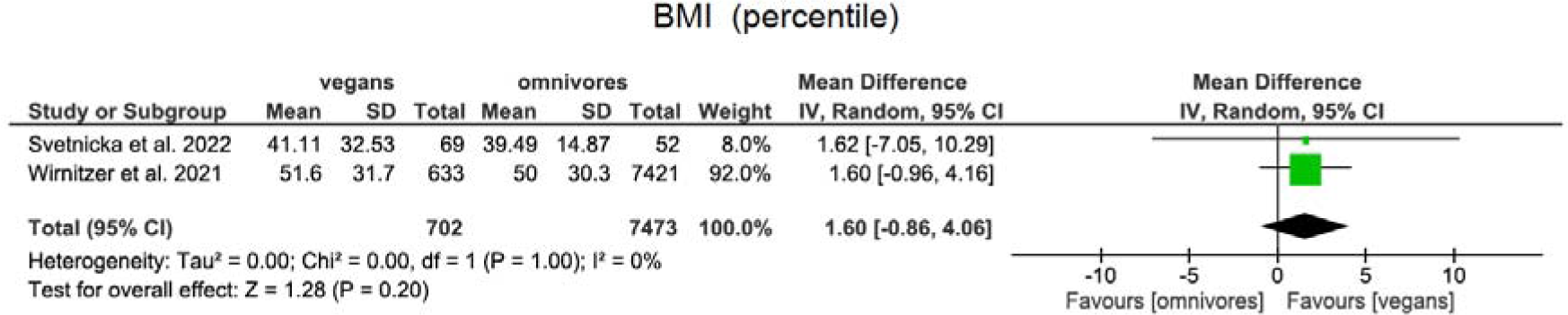
Mean differences with 95CIs using a random effects model for a vegan diet compared to an omnivore diet regarding BMI in percentiles among children.

#### Single study findings concerning anthropometric values

Length after 12 months, weight 6 and 12 months (37) fat mass index, suprailiac skinfold, triceps skinfold, tight girth, as well as hip girth (33) were lower among vegans compared to omnivores.

Birth length, length after 6 months (37), weight for height, weight for age (31), lean mass index, biceps skinfold thickness, subscapular skinfold thickness, waist girth (33), head circumference at birth, after 6 months, after 12 months (37) showed no difference between vegans and omnivores.

#### Biomarkers of metabolism / miscellaneous

All outcomes concerning biomarkers are presented in Table 4.

**Table 4.**
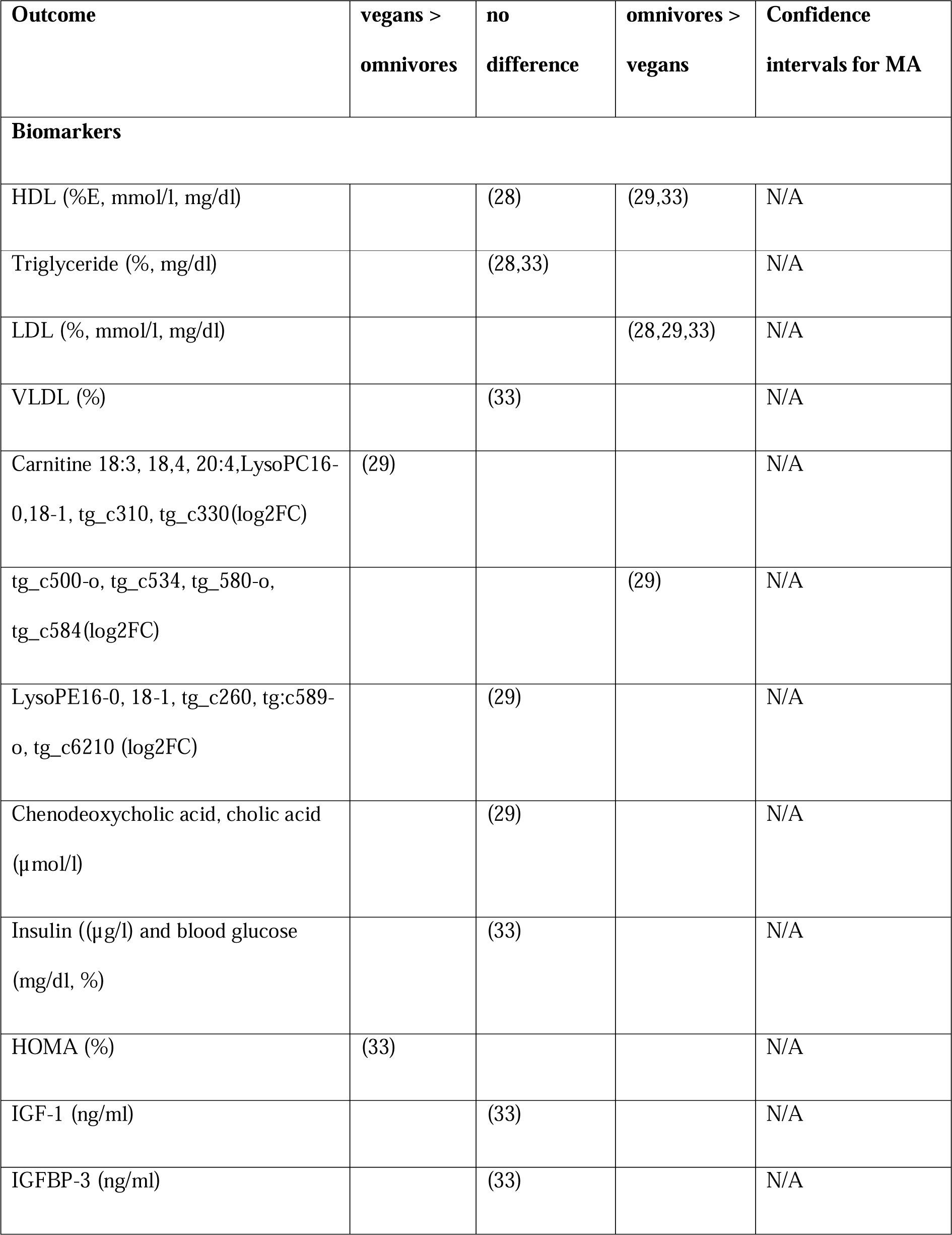
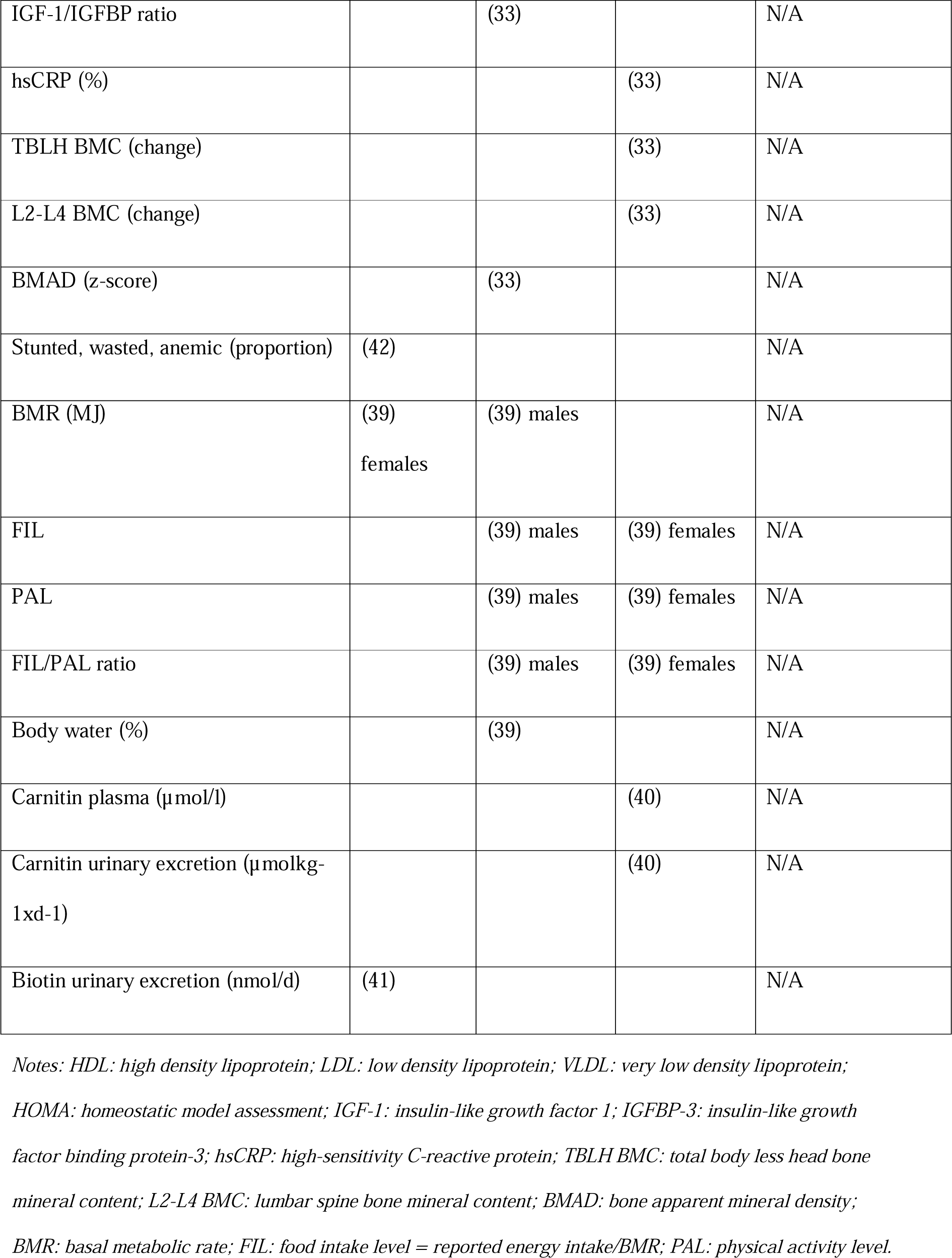
Overview of biomarker differences in vegan compared to omnivorous children.

| Outcome | vegans ><br>omnivores | no<br>difference | omnivores ><br>vegans | Confidence<br>intervals for MA |
| --- | --- | --- | --- | --- |
| **Biomarkers** |  |  |  |  |
| HDL (%E, mmol/l, mg/dl) |  | (28) | (29,33) | N/A |
| Triglyceride (% , mg/dl) |  | (28,33) |  | N/A |
| LDL (% , mmol/l, mg/dl) |  |  | (28,29,33) | N/A |
| VLDL (%) |  | (33) |  | N/A |
| Carnitine 18:3, 18:4, 20:4, LysoPC16-0, 18-1, tg_c310, tg_c330(log2FC) | (29) |  |  | N/A |
| tg_c500-o, tg_c534, tg_580-o, tg_c584(log2FC) |  |  | (29) | N/A |
| LysoPE16-0, 18-1, tg_c260, tg:c589-o, tg_c6210 (log2FC) |  | (29) |  | N/A |
| Chenodeoxycholic acid, cholic acid (μmol/l) |  | (29) |  | N/A |
| Insulin ((μg/l) and blood glucose (mg/dl, %) |  | (33) |  | N/A |
| HOMA (%) | (33) |  |  | N/A |
| IGF-1 (ng/ml) |  | (33) |  | N/A |
| IGFBP-3 (ng/ml) |  | (33) |  | N/A |
| IGF-1/IGFBP ratio |  | (33) |  | N/A |
| hsCRP (%) |  |  | (33) | N/A |
| TBLH BMC (change) |  |  | (33) | N/A |
| L2-L4 BMC (change) |  |  | (33) | N/A |
| BMAD (z-score) |  | (33) |  | N/A |
| Stunted, wasted, anemic (proportion) | (42) |  |  | N/A |
| BMR (MJ) | (39)<br>females | (39) males |  | N/A |
| FIL |  | (39) males | (39) females | N/A |
| PAL |  | (39) males | (39) females | N/A |
| FIL/PAL ratio |  | (39) males | (39) females | N/A |
| Body water (%) |  | (39) |  | N/A |
| Carnitin plasma ($\mu\text{mol/l}$) | | | (40) | N/A |
| Carnitin urinary excretion ($\mu\text{molkg}^{-1}\text{d}^{-1}$) | | | (40) | N/A |
| Biotin urinary excretion (nmol/d) | (41) |  |  | N/A |

Notes: HDL: high density lipoprotein; LDL: low density lipoprotein; VLDL: very low density lipoprotein;
HOMA: homeostatic model assessment; IGF-1: insulin-like growth factor 1; IGFBP-3: insulin-like growth
factor binding protein-3; hsCRP: high-sensitivity C-reactive protein; TBLH BMC: total body less head bone
mineral content; L2-L4 BMC: lumbar spine bone mineral content; BMAD: bone apparent mineral density;
BMR: basal metabolic rate; FIL: food intake level = reported energy intake/BMR; PAL: physical activity level.

Three studies examined cardiovascular risk markers. Two of three studies (29,33) showed lower HDL (high density lipoprotein) levels in vegans. Two studies (28,33) showed no difference in triglyceride levels. All three studies (28,29,33) showed lower LDL (low density lipoprotein) levels in vegans.

#### Single study findings concerning metabolism / miscellaneous

Red blood count (RBC), hemoglobin (Hb), platelets (28), CRP, total body less head bone mineral content (TBLH BMC), the lumbar spine bone mineral content (L2-L4 BMC) (33), basal metabolic rate (BMR (only in girls)), food intake level (FIL), physical activity level (PAL) (only in girls) (39), carnitine in plasma (µmol/l) and urinary carnitine excretion (µmolkg-1xd-1) (40) were lower in vegans compared to omnivores.

Biotin excretion (41), proportion wasted, stunted, anemic (42) and HOMA levels (35), MCV with and without supplementation (33) were higher in vegans compared to omnivores.

The mean corpuscular volume (MCV) (34), blood glucose, insulin, VLDL, IIGF-1 (insulin-like growth factor 1), IGFBP-3 (insulin-like growth factor binding protein-3), Molar IGF-1/IGFBP ratio (33), food intake level (FIL=reported energy intake/BMR (basal metabolic rate), PAL (only in males), their ratio (FIL/PAL), BMR (only in males) and body water content (39) showed no difference between vegans and omnivores.

#### Randomized controlled trial

A randomized controlled trial (RCTs) (43) by Macknin et al. examined children whose BMI was above the 95th BMI percentile and cholesterol levels above 169mg/dl. The intervention was either a vegan diet, a diet recommended by the American Heart Association (AHA diet) or a and a Mediterranean diet. The group of vegan children showed a greater reduction in protein, vitamin B12 and vitamin D intake. In all three groups, cholesterol intake, waist circumference, fasting glucose, systolic and diastolic blood pressure, and myeloperoxidase (MPO) decreased. There was no reduction in BMI, HDL cholesterol, insulin level and hs-CRP in all three groups. Physical activity levels changed only in the Mediterranean diet group.

## Discussion

Our systematic review (SR) identified 17 primary studies 16 cross-sectional studies and one randomized controlled trial) on health outcomes among vegan children. The outcomes were mainly nutrient intakes, biomarkers of nutrients status, anthropometric parameters and metabolic biomarkers.

Our meta-analyzes showed higher carbohydrate, fiber and PUFA intakes in vegans, lower intakes of protein and SFA in vegans. No difference was found in total fat, MUFA intake, birth weight and BMI between vegans and omnivores.

A wide range of further outcomes were not meta-analyzable, but individual studies showed the following results: Vegan children were found to have lower intakes of docosahexaenoic acid, eicosapentaenoic acid, vitamin B2, vitamin B12, zinc and selenium as well as lower blood concentrations of ferritin and vitamin B12. On the other hand, vegan children had higher intakes of folate, vitamin C, vitamin B1, vitamin B6, magnesium, iron and alpha-linolenic acid. Levels of different cardiometabolic risk markers (total cholesterol, LDL, blood pressure) were lower in vegans. Risk factors and indicators of bone health (vitamin D and calcium intake, vitamin D blood levels, as well as bone mineral density) were lower in vegans. No differences were observed regarding vitamin A/beta carotene intake, blood insulin, CRP and IGF levels, as well as triglyceride levels.

The risk of bias in primary studies was graded high or very high in 7 out of 17 primary studies using ROBINS-E/ ROB 2.0. The certainty of evidence was rated very low or low for all nine included meta-analyses using GRADE.

### Specific findings

#### Protein intake

Our meta-analysis of three primary studies showed a lower relative protein intake in vegan children although two further studies revealed no significant difference in absolute protein intake. One study included in the meta-analysis showed ranges for the recommended daily protein intake (%E). Although intakes were lower in vegans than in omnivores, both groups achieved the recommended daily intake (30). Of note, only one study compared the total energy density, which was significantly lower in vegans, and may explain lower total protein intakes (28). However, the lower relative protein intake cannot be explained by this finding. Besides protein quantity, protein quality may be lower among vegans. While a combination of various sources of plant protein (vegetables, legumes, grains, nuts) may facilitate sufficient protein quality and bioavailability (44). On the one hand children do have higher protein requirements per kg of body weight than adults (45). On the other hand, expressing these requirements as percentage of energy, the requirements are substantially lower than for adults (44). Therefore, as children have an increased energy demand compared to adults, meeting protein requirements on a vegan diet is easily possible, if the diet does not exceed reasonable amounts of refined oils and sugars.

#### Fat intake

The meta-analyses on relative fat intake showed no difference between vegan and omnivore children, while saturated fat intake was lower and unsaturated fat intake was higher among vegans compared with omnivores.

With respect to n-3 fatty acids, our SR and previous studies among adults showed that the supply with plant-derived ALA is better among vegans, while the status of animal-derived (mainly fish) n-3 fatty acids EPA and DHA is lower (46). ALA can be metabolized into EPA and DHA, albeit at a low conversion rate (46,47). The conversion rate can be reduced by a high intake of n-6 fatty acids, but is vital in a vegan diet, because EPA and DHA are not found in regular plant products (except some microalgae), consumed in a vegan diet. EPA and DHA may have anti-inflammatory and cardioprotective properties (47) and may have a beneficial impact on cognitive function (48). In our SR, vegan children had a lower intake of EPA and DHA but a higher intake of ALA.

A study by Sanders et al. (49) examined polyunsaturated fatty acids in the erythrocytes of newborns breastfed by vegan/omnivore mothers. 20:3n-6 and 22:4n-6 was significantly higher in vegans, but DHA was significantly lower. As this study was of very small sample size (three vegans vs six omnivores), more research is needed to draw conclusions about the potential inadequacy of effective n-3 fatty acid supply and the adequacy of the conversion rate from ALA to DHA and EPA. In the future supplementation with algae oil (containing DHA and EPA) may be considered for providing potential deficiencies in children.

Concerning n-6 fatty acids, intake of AA as well as AA concentration in erythrocytes was lower in vegan children, but the intake and concentration in erythrocytes of LA was higher in vegan children. The lower intake AA may be explained by the low content of arachidonic acid in plant foods and high levels in meat, especially sausage products (50). Considering the pro-inflammatory properties of n-6 fatty acids, especially AA, vegan children seem to have a more favorable profile of these fatty acids (48).

#### Fiber

Our SR showed a higher relative intake of dietary fiber in vegans than in omnivore children. Studies conducted in adults support these findings (51,52). Adequate fiber intake has been associated with a generally favorable pattern of metabolic biomarkers and lower risks of several chronic diseases (53). For example, fiber intake is related to lower cholesterol levels (54). Fiber is also associated with improved insulin sensitivity and thus a reduced risk of type 2 diabetes mellitus, which may be of importance due to the increasing prevalence of type 2 diabetes in children (55). Many of the health benefits of dietary fiber are mediated via the composition and function of the intestinal microbiome (56). Fiber intake also leads to increased intestinal motility and softer stool consistency, thereby potentially reducing the risk of hemorrhoids and diverticula (53).

#### Vitamin B12

Two of the studies that we included differentiated between vitamin B12 intake with and without supplementation, showing a lower intake without supplements in vegans. Among vegan children who took vitamin B12 supplements, total intakes were higher compared to omnivore children. Vitamin B12 has an important role in cell metabolism, especially in DNA synthesis (57). Two studies compared the intake with ranges for the recommended daily vitamin B12 intake (µg/d). Vegans did not reach the recommended daily intake in both studies, if not supplemented (30, 31). In adults, vitamin B12 deficiency can manifest with neurological, neuropsychological, or hematological symptoms (normochromic macrocytic anemia). In children, such symptoms have been rarely reported. Some case studies showed developmental delay in children with vitamin B12 deficiency (58). There is no known way to increase the absorption of vitamin B12 in contrast to the other nutrients identified as critical in this review. Additionally, no relevant amounts of vitamin B12 are found in plant-based foods, therefore supplementation is essential and should be strongly recommended by treating physicians.

#### Zinc

Although in our review zinc intake was lower among vegans in only one out of four studies (30) and the same study showed, that vegans reached the recommended daily intake, possible risks of zinc deficiency among vegans are being discussed. As a recent systematic review from 2021 showed significantly lower zinc intakes in vegan adults (59). Good plant sources of zinc include legumes, grains, and various vegetables, but zinc absorption from plant foods can be reduced by high phytate contents (60). Zinc serves as an important cofactor in various enzymes in the body and thus has catalytic and regulatory functions (61). Zinc deficiency in childhood can lead to growth retardation, immunodeficiencies, gastrointestinal and dermatologic issues (62). However, blood zinc levels were determined in only one study (29) in our SR, and were not significantly lower among vegan children. Thus, further studies on zinc status among vegan children are needed.

#### Iodine

In our review, iodine intake was only assessed in a single study and was lower in vegans (31). In a second study, no differences in the urinary iodine to creatinine ratio between vegans and omnivores were observed (29). Despite the few studies among vegan children, possible risks of an iodine deficiency among vegans are being discussed. Previous studies indicate possible iodine deficiencies among vegan adults (63). A systematic review from 2020 among adults stated a lower iodine intake in vegans. Iodine is essential for the synthesis of thyroid hormones T3 (triiodothyronine) and T4 (thyroxin). These hormones are crucial for the regulation of the metabolism and other processes such as growth and neurological development. Low iodine intake in mothers during pregnancy and lactation is known to cause congenital hypothyroidism (cretinism i.e. mental and physical retardation due to an untreated congenital hypothyroidism) in infants (64).

As our SR provides only little data concerning iodine intake, but potential deficiencies may be of crucial importance for the development of vegan children, further research is needed.

#### Selenium

Selenium intake was assessed in two studies and was lower in vegans compared to omnivores (30,35). In one study the average requirement of 17 µg/d was reached in both groups (35), but in the other the recommended intake was not reached in vegans but in omnivores (30). A previous systematic review showed a lower intake of selenium among vegan adults compared to omnivores (59). Selenium is physiologically included in many processes as thyroid hormone production, proper immune system functioning and works as an anti-oxidant (65). During pregnancy selenium intake may be essential for neurodevelopment. It has been proposed that lower selenium status during pregnancy may correlate with higher risks of autism spectrum disorder (ASD) and attention deficit hyperactivity disorder (ADHD)(66). The results in our review do not show deficiencies but only lower levels in vegans compared to omnivores. As deficiencies, especially during neurodevelopment may be of serious consequences, further research is needed.

#### Bone health

Vitamin D and calcium intake, which are essential for bone health and prevention of osteopenia, were lower in vegan children. One of the included studies showed that vegans in contrast to omnivores did not reach the recommended daily intake of calcium (30). A single study by Desmond et al. (33) compared BMC of the lumbar spine and total body between vegan and omnivore children. Both were lower in the vegan group, but no difference in apparent bone mineral density was found (33). Although these results are from a single study with a very small sample size (52 vegan vs. 72 omnivore children), bone health of vegan children may be considered as critical due to the equally lower calcium and vitamin D intake values. These findings may be relevant in the context of higher fracture risk that has been described among adult vegans (12). It is important to mention that peak bone mass is formed in adolescence up to about 25 years of age. If the intake of vitamins and minerals important for bone metabolism is reduced, the maximum peak bone mass might be lower in vegans. This would increase the risk of fractures in adults and due to the lower peak bone mass and osteoporosis may manifest earlier (67). It is therefore of great importance to pay attention to vitamin and mineral intake (vitamin D3, calcium) in a vegan diet. Good sources of calcium for vegan children are mineral water, calcium enriched plant-based milk alternatives, tofu, sesame and green leafy vegetables especially cruciferous vegetables (68). The bioavailability of these products may differ a lot due to the high content of oxalic acid, which can be reduced by steaming or boiling the vegetables (69). Alternatively, food can be enriched with calcium (from corals/ algae). Vitamin D sources, however, are limited in the vegan diet (70). Thus, a supplementation with vitamin D can be considered, especially in the winter months.

Besides sufficient nutrient intake, sufficient physical activity during growth is essential for bone modeling. Out of the studies identified for this SR, only the one by Larsson et al. (39) reported data on physical activity, but no difference between vegan and omnivore children was found. In the long-term, follow-up studies of adults/ seniors who have been on a vegan diet since childhood would be desirable to accurately compare the risk of osteoporosis/fracture (especially regarding the potentially reduced peak bone mass).

#### Cardiometabolic risk markers

Concerning cardiometabolic risk factors, no meta-analysis could be performed due to the different age groups and various biomarkers with mixed units (relative/absolute). Nevertheless, lower LDL and total cholesterol levels among vegan children in the identified studies indicate a lower cardiovascular risk profile. Previous studies among adults suggested a lower 10-year CHD risk in adult vegans with hyperlipidemia and lower mortality due to cardiovascular events in vegans (12). Whether a more favorable cardiometabolic risk profile in childhood leads to an additional risk reduction in adulthood among long-term vegans needs investigation in life course studies.

#### Risk of anemia

One study showed a higher prevalence of anemic children for mothers consuming a vegan compared to an omnivorous diet (41). The study was conducted in India and is therefore probably not directly applicable to Western countries. Nevertheless, a study observed lower hemoglobin levels in vegans in Germany (18). Additionally, two out of three studies showed lower ferritin levels among vegan compared to omnivore children(28,33) may point to an increased anemia risk among vegan children and further studies in ethnically and culturally diverse populations are needed.

Total iron intake is often higher in vegans but ferritin levels may be lower. This finding can be explained by the fact that the bioavailability of iron from plant-foods is significantly lower than from animal products, in part due to the high content of phytic acid, which acts as an inhibiting factor in iron absorption. However, the bioavailability of iron from plant foods can be increased to a certain extent by the additional intake of foods rich in vitamin C (71). Possible consequences of anemia in childhood include impaired cognitive and physical development and immune system, as well as increased risk of infectious diseases and overall mortality (72). Therefore, a sufficient supply of iron among vegan children should be monitored.

## Strength and limitations

This SR has several strengths. It is the first systematic review summarizing and evaluating all publications on health outcomes among vegan children and adolescents. We followed state-of-the art procedures (pre-registered protocol, reporting guidelines) and we examined the risk of bias and certainty of the evidence using validated tools (ROBINS-E, GRADE).

However, this SR has also several limitations. High or very high risk of bias was detected in seven out of 17 primary studies. In many studies, sample sizes were small and no adjustment for essential confounders was carried out. Most studies only compared the outcomes of vegan children to omnivores and not to recommended daily intakes/ reference values. This would be helpful to differentiate between outcome values among vegan children which are “lower than among omnivores” vs. not reaching recommended reference levels, and therefore risking potential deficiencies. In addition, some of the included studies were rather old, and it is not clear whether dietary habits in these older studies are reflective of dietary habits today. Further, most studies did compare nutrient intakes or blood biomarkers, which may indicate negative effects on the development of children, but no studies were found concerning “hard outcomes” such as cognitive or motor function or fracture risk/ risk of osteoporosis in later stages of life were available. Studies on questionnaire-derived dietary intakes may have generally underestimated nutrient intakes (73). A methodological difficulty when investigating studies among children and adolescents is the limited comparability of certain outcomes across different age groups, especially as no age adjustment was used for many studies in this SR.

## Conclusion

As veganism is gaining in popularity, high-quality evidence is needed to guide vegan families and their practitioners. Overall, our findings on health outcomes among vegan children are well in line with findings from previous studies among adults. Vegan children may have lower intakes of energy, protein, and lower vitamin B12 levels if no supplements are taken. Especially studies on protein quantity and quality are needed. Calcium and vitamin D intake, vitamin D blood concentrations, and BMD may be lower in vegans. While vegan children may be less exposed to pro-inflammatory n-6 fatty acids, intakes of essential n-3 fatty acids, EPA and DHA, are lower in a vegan diet. Future studies are needed on sources and bioavailability of these n-3 fatty acids among vegans. Cardiometabolic risk profiles among vegan children seem generally favorable compared with omnivore children. Generally, future studies should compare dietary intakes and biomarker levels to reference ranges, as the mere observation of statistically significant mean differences in certain parameters between vegan and non-vegan children as such may not be informative with regard to deficiency risks or other health outcomes. In clinical practice, monitoring critical nutrients or biomarkers in blood testing has to be weighed individually against the best interest of the child, as taking blood samples is an invasive intervention. An alternative to standard testing for critical nutrients would be to seek advice from a qualified nutritionist who can assess the need for monitoring on an individual basis. Certain noninvasive monitoring options for example urine samples (for iodine) or sonography for bone mineral density may be considered.

## Disclamer

This work reflects only the authors’ view. The Swiss Federal Office of Public Health is not responsible for any use that may be made of the information it contains.

## Author Contributors

AK, JK and TK designed the research. AK, JK and TK conducted the literature search and literature screening, extracted the data. JK analyzed and recalculated the data. AK and JK assessed the risk of bias and the certainty of evidence. AK wrote the first draft of the paper, JK and TK revised it. SR, MW, JG, ES, MC, SS, TK, JK, MS, JK interpreted the data, read the manuscript and approved the final version.

## Supporting information

file:///Users/familiekoller/Desktop/Supplementary%20appendix.html

## Data Availability

All data produced in the present work are contained in the manuscript

