## Supplementary material for "Health aspects of vegan diets among children and adolescents: a systematic review and meta-analyses": file:///Users/familiekoller/Desktop/Supplementary%20appendix.html: Supplementary appendix.pdf

Table S1. Search strategy for pubmed

| | |
|---|---|
| *# 1* | "vegans"[MeSH Terms] OR "diet, vegan"[MeSH Terms] OR vegan* OR ("plant-based" AND(diet* OR food* OR nutri*)) |
| *# 2* | "child"[MeSH Terms] OR child* OR "adolescent"[MeSH Terms] OR adolesc* OR "infant"[MeSH Terms] OR infant* OR toddler* OR juvenile OR youth |
| *# 3* | *Combine: # 1 AND # 2* |

Table S2. Search strategy for EMBASE

| | |
|---|---|
| *# 1* | ('vegan'/exp OR 'vegan' OR 'vegan diet'/exp OR 'vegan diet' OR vegan* OR ('plant-based' AND (diet* OR food* OR nutri*))) |
| *# 2* | ('child'/exp OR 'child' OR child* OR 'adolescent'/exp OR 'adolescent' OR adolesc* OR adolescen* OR 'juvenile'/exp OR 'juvenile' OR 'infant'/exp OR 'infant' OR infant* OR toddler* OR 'toddler'/exp OR 'toddler' OR 'youth'/exp OR 'youth') |
| *# 3* | *Combine: # 1 AND # 2* |

Table S3: PICOS statement summarizing study rationale and study selection criteria

| | **PICOS Statement** | |
|---|---|---|
| **P** | Population | Children and adolescents 0 to 18 years of age |
| **I** | Intervention/ Exposure | Vegan diet: defined as a plant-based diet avoiding any product of animal origin (except human breastmilk) for at least one year/ since birth |
| **C** | Comparison | Omnivore diet: consuming all types of foods, including products of animal origin |
| **O** | Outcome | Any health outcome and nutritional status |
| **S** | Study Design | Cross-sectional studies, Randomized controlled trials, prospective studies |

Figure S1: Flow chart of the study selection progress

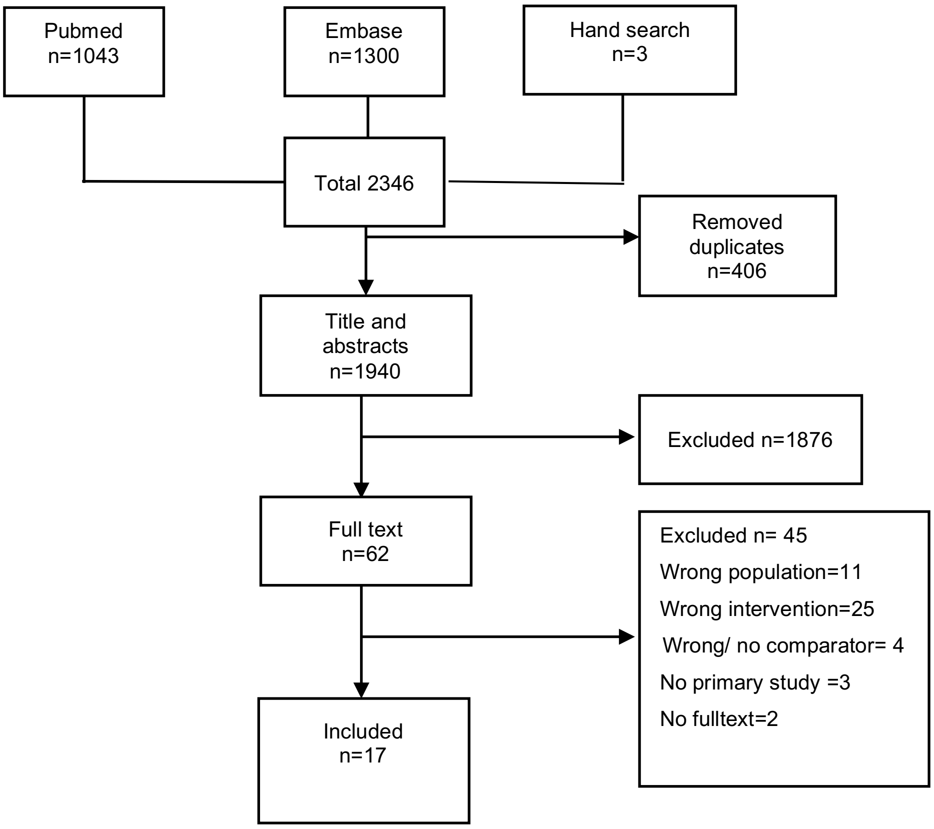

*Table S4:* List of excluded studies

| **Reasons for exclusion** | **Reference** |
|---|---|
| Wrong population | 11 (1–11) |
| Wrong intervention | 24 (12–36) |
| No Comparison | 4 ((37–40) |
| No fulltext | 2 (41,42) |
| No primary study | 3 (43–45) |

Figure S2: Correlation plot between effect size per outcome and age

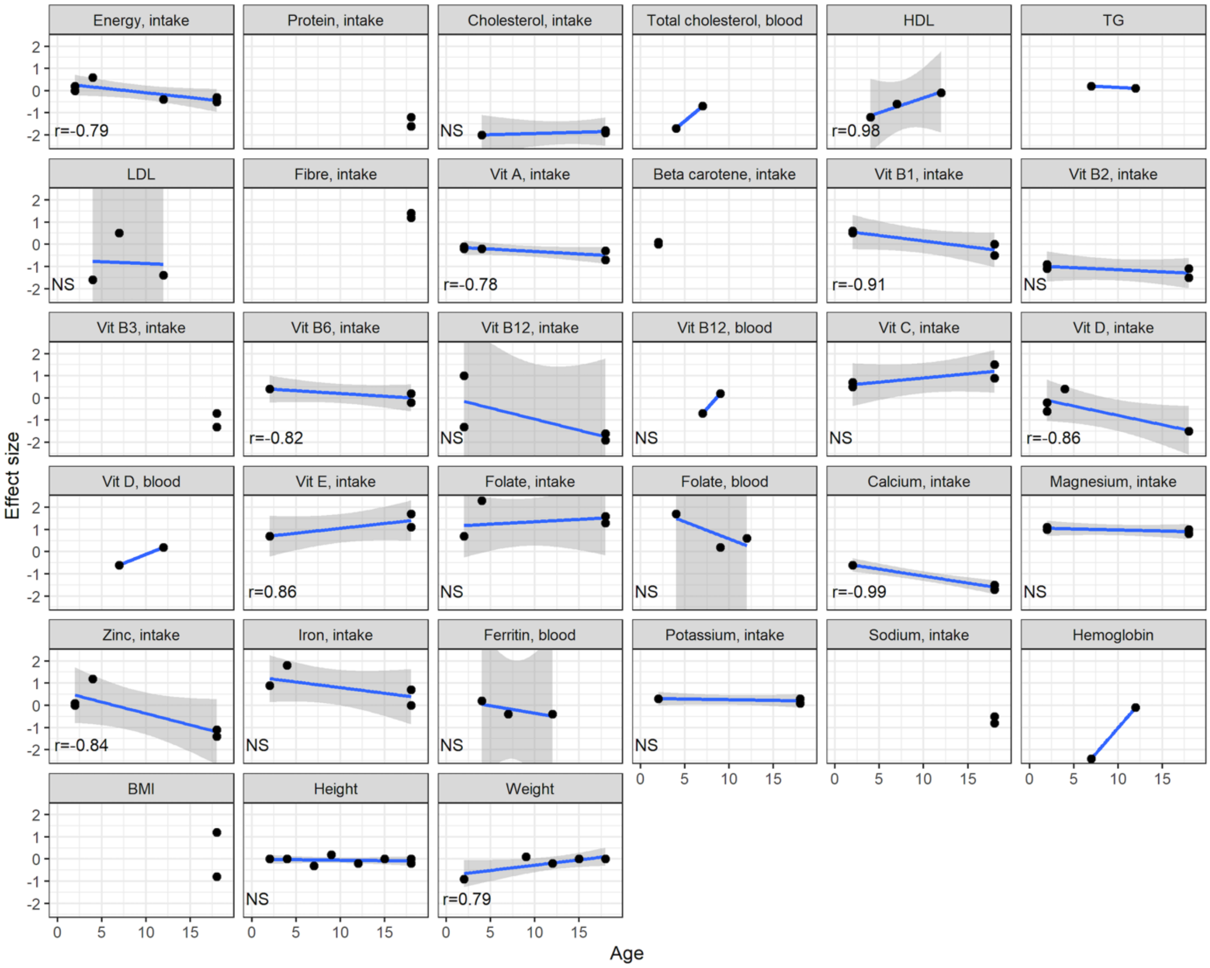

Table S5: Characteristics of the included studies on a vegan diet regarding age, country, sample size, adjusted confounders and risk of bias.

| Nr | Author (Year)[1] | Age | Country | n (v/o) | Adjusted confounders |
|---|---|---|---|---|---|
| 1 | Alexy et al. (2021) | 6 to 18 Y | Germany | 114/137 | age, sex, suppl., SES, PA, BMI, puberty, parental smoking |
| 2 | Desmond et al. (2021) | 5 to 10 Y | Poland | 52/72 | age, sex, suppl., edu., PA, BMI, parental smoking |
| 3 | Ferrara et al (2021) | 0 to 12 M | Italy | 21/21 | none |
| 4 | Headey et al. (2020) | 24 to 59 M | India | unclear/r.v. | age, sex, SES |
| 5 | Hovinen et al. (2021) | median 3.5 Y | Finland | 6/24 | age, sex |
| 6 | Larsson et al. 1, m (2002) | 17.4+-0.8 Y | Sweden | 9/9 | age, sex, height |
| 6 | Larsson et al. 1, f (2002) | 17.4+-0.8 Y | Sweden | 7/7 | age, sex, height |
| 7 | Larsson et al. 2, m (2002) | 17.5+-1 Y | Sweden | 15/15 | age, sex, height |
| 7 | Larsson et al. 2, f (2002) | 17.5+-1 Y | Sweden | 15/15 | age, sex, height |
| 8 | Lombard et al. (1989) | 2.4 to 17.7 Y | USA | 25/29 | sex, suppl. |
| 9 | Lombard et al. (1989) | 2.4 to 15.1 Y | USA | 25/29 | suppl. |
| 10 | Macknin et al. (2021) | 9 to 18 Y | USA | 25/27 | age, sex |
| 11 | Pawlak et al. (2014) | newborns | USA | 47/350 | none |

| | | | | | |
|---|---|---|---|---|---|
| 12 | Sanders et al. 1(1992) | 14 W | UK | 3/6 | gestational age, sex, parity |
| 13 | Svetnicka et al. (2022) | 0 to 18 Y | Czech republic | 69/52 | age, sex, suppl., BMI |
| 14 | Weder et al. (2019) | 1 to 3 Y | Germany | 139/164 | age, sex, suppl., SES, PA, BMI par., urb. |
| 15 | Weder et al. (2021) | 1 to 3 Y | Germany | 139/164 | age, sex, suppl., SES, PA, BMI, puberty, urb. |
| 16 | Weder et al. (2023) | 1 to 3 Y | Germany | 139/164 | age, sex, suppl., SES, PA, BMI, puberty, urb. |
| 17 | Wirnitzer et al. (2021) | 15.1+-2.3 Y | Austria | 633/7421 | age, sex, BMI, PA |

Abbreviations: Y: years; M: months; n: sample size; v: vegan; o: omnivore, suppl.: supplements, SES: socioeconomic status, PA: physical activity, edu.: parental education, urb.: urbanicity, RoB: risk of bias

RoB was assessed using ROBINS-E and RoB2.0. for randomized controlled trials.
All studies except one randomized controlled study (Macknin) are cross-sectional studies.

Table S6: Results of all outcomes of the included 19 studies

vgn, vegan; omni; omnivores; SD, standard deviation

| *Author* | *Outcome* | *Unit* | *n vgn* | *Mean (SD) vgn* | *n omni* | *Mean (SD) omni* | *Difference (vgn-omni)* | *CIs* |
|---|---|---|---|---|---|---|---|---|
| **Energy intake** | | | | | | | | |
| Alexy et al. (2021) | energy | kcal/day | 110 | 1634.3(405.5) | 135 | 1859.6(584.5) | -225.3 | [-350.3;-100.3] |
| Alexy et al. (2021) | energy density | (kj/g)3 | 110 | 5.4(1.1) | 135 | 6.3(1.5) | -0.9 | [-1.3;-0.6] |
| Hovinen et al. (2021) | energy | kj/d | 6 | 5841.8(790.8) | 24 | 5208.7(1036.5) | 633.1 | [-240.1;1506.2] |
| Weder et al. (2021) | energy | MJ/d | 139 | 4.1(0.45047) | 164 | 4.1(1) | 0.0 | |
| Weder et al. (2019) | energy | kcal/d | 139 | 1026.4(280) | 164 | 986.2(181.5) | 40.2 | [-14.2;94.7] |
| Weder et al. (2019) | energy | kcal/g | 139 | 1.1(0.2) | 164 | 1.2(0.2) | -0.1 | [-0.2;-0.1] |
| Larsson et al. 2, m (2002) | energy | MJ | 15 | 12.2(1.7) | 15 | 13.2(2) | -1.0 | [107.4;110.2] |
| Larsson et al. 2, f (2002) | energy | MJ | 15 | 8.9(2.2) | 15 | 9.5(2) | -0.6 | [-2.1;1] |
| **Alcohol** | | | | | | | | |
| Larsson et al. 2, m (2002) | alcohol | g | 15 | 2.1(3.1) | 15 | 7(7.9) | -4.9 | [-9.5;-0.3] |
| Larsson et al. 2, m (2002) | alcohol | %E | 15 | 0.5(0.7) | 15 | 1.6(1.8) | -1.1 | [-2.1;-0.1] |
| Larsson et al. 2, f (2002) | alcohol | g | 15 | 3.9(4.4) | 15 | 4.9(7) | -1.0 | [-5.4;3.4] |
| Larsson et al. 2, f (2002) | alcohol | %E | 15 | 1.4(1.6) | 15 | 1.6(2.3) | -0.2 | [-1.7;1.3] |
| **Sugar** | | | | | | | | |
| Larsson et al. 2, m (2002) | disaccharides | g | 15 | 77(20) | 15 | 137(40) | -60.0 | [-84.1;-35.9] |
| Larsson et al. 2, f (2002) | disaccharides | g | 15 | 72(29) | 15 | 100(29) | -28.0 | [-49.7;-6.3] |

| | | | | | | | | |
|---|---|---|---|---|---|---|---|---|
| Alexy et al. (2021) | free sugars | %E | 110 | 6.7(3.8) | 135 | 12.5(7.8) | -5.8 | [-7.3;-4.3] |
| Larsson et al. 2, m (2002) | monosaccharides | g | 15 | 64(19) | 15 | 47(17) | 17.0 | [3.5;30.5] |
| Larsson et al. 2, f (2002) | monosaccharides | g | 15 | 65(28) | 15 | 48(16) | 17.0 | [-0.3;34.3] |
| Larsson et al. 2, m (2002) | sucrose | g | 15 | 73(21) | 15 | 88(36) | -15.0 | [-37.3;7.3] |
| Larsson et al. 2, f (2002) | sucrose | g | 15 | 69(28) | 15 | 69(22) | 0.0 | [-18.9;18.9] |
| Weder et al. (2019) | added sugars | %E | 139 | 4.3(6.8) | 164 | 5.9(5) | -1.6 | [-3;-0.3] |
| **Fibre** | | | | | | | | |
| Alexy et al. (2021) | fiber | g/1000kcal | 110 | 21.2(5.8) | 135 | 12.6(3.2) | 8.5 | [7.3;9.7] |
| Weder et al. (2019) | fiber | g/1000kcal | 139 | 20.7(6.2) | 164 | 13.2(4.6) | 7.5 | [6.2;8.7] |
| Larsson et al. 2, m (2002) | fiber | g | 15 | 44(10) | 15 | 25(8) | 19.0 | [12.2;25.8] |
| Larsson et al. 2, f (2002) | fiber | g | 15 | 34(11) | 15 | 21(6.3) | 13.0 | [6.2;19.8] |
| **Protein intake** | | | | | | | | |
| Alexy et al. (2021) | protein | g/kg BW/d | 110 | 1.3(0.7) | 135 | 1.4(0.5) | -0.1 | [-0.3;0.1] |
| Weder et al. (2021) | protein | g/kg BW | 139 | 2.3(0.9) | 164 | 2.5(0.9) | -0.3 | |
| Larsson et al. 2, m (2002) | protein | g | 15 | 73(13) | 15 | 117(21) | -44.0 | [-57.2;-30.8] |
| **Protein blood** | | | | | | | | |
| Lombard et al. 1 (1989) | protein in serum | g/l | 25 | 43.3(3.2) | 29 | 74(4.9) | -30.7 | [-32.9;-28.5] |
| **Carbohydrate intake** | | | | | | | | |

| | | | | | | | | |
|---|---|---|---|---|---|---|---|---|
| Larsson et al. 2, m (2002) | carbohydrates | g | 15 | 456(62) | 15 | 424(75) | 32.0 | [-19.6;83.6] |
| Larsson et al. 2, m (2002) | carbohydrates | g | 15 | 340(79) | 15 | 310(62) | 30.0 | [-23.3;83.3] |
| **Fat intake** | | | | | | | | |
| Larsson et al. 2, m (2002) | fat | g | 15 | 25(5.5) | 15 | 45(9.3) | -20.0 | [-25.8;-14.2] |
| Larsson et al. 2, f (2002) | fat | g | 15 | 15(6.8) | 15 | 34(10) | -19.0 | [-25.4;-12.6] |
| Larsson et al. 2, m (2002) | fat | g | 15 | 88(15) | 15 | 100(22) | -12.0 | [-26.2;2.2] |
| Larsson et al. 2, f (2002) | fat | g | 15 | 58(23) | 15 | 75(22) | -17.0 | [-33.9;-0.1] |
| **n-3/ n-6 fatty acids** | | | | | | | | |
| Weder et al. (2021) | 18:2,-6(LA) | E% | 139 | 7.2(1.6) | 164 | 3.9(1.3) | 3.3 | [3;3.7] |
| Weder et al. (2021) | 18:3n-3(ALA) | E% | 139 | 1.6(1.5) | 164 | 0.6(0.3) | 1.0 | [0.8;1.3] |
| Weder et al. (2021) | 20:4n-6(AA) | mg/d | 139 | 12.6(14.9) | 164 | 44(37.6) | -31.4 | [-37.7;-25.1] |
| Weder et al. (2021) | 20:5n-3(EPA) | mg/d | 139 | 5(3.6) | 164 | | | |
| Weder et al. (2021) | 20:5n-3(EPA) | mg/d | 139 | 4.8(3.8) | 164 | 31.5(7.7) | -26.7 | [-28.1;-25.4] |
| Weder et al. (2021) | 22:6n-3(DHA) | mg/d | 139 | 28.5(28.8) | 164 | 81.1(146.2) | -52.6 | [-75.7;-29.6] |
| Weder et al. (2021) | 22:6n-3(DHA) | mg/d | 139 | 24.3(19.9) | 164 | 72.2(123.1) | -47.9 | [-67.2;-28.7] |
| Weder et al. (2021) | LA:ALA | | 139 | 6.8(3.3) | 164 | 6.3(2.2) | 0.5 | [-0.1;1.2] |
| Hovinen et al. (2021) | ALA | log2 FC | 6 | 1.5(0.4) | 24 | 1(0.5) | 0.5 | [0;0.9] |
| Hovinen et al. (2021) | DHA | log2 FC | 6 | 0.5(0.1) | 24 | 1(0.4) | -0.5 | [-0.7;-0.3] |
| **Fatty acids (erythrocytes)** | | | | | | | | |
| Sanders et al.1 (1992) | 18:2n-6 | | 3 | 10.9(2.7) | 6 | 6.6(0.3) | 4.3 | [-2.4;11] |

| | | | | | | | | |
|---|---|---|---|---|---|---|---|---|
| Sanders et al.1 (1992) | 20:3n-6 | | 3 | 1.3(0.1) | 6 | 0.9(0.1) | 0.4 | |
| Sanders et al.1 (1992) | 20:4n-6(AA) | | 3 | 13.3(0.6) | 6 | 13.7(0.6) | -0.4 | [0.1;0.7] |
| Sanders et al.1 (1992) | 20:5n-3(EPA) | | 3 | 0.2(0) | 6 | 0.7(0.6) | -0.5 | [-1.6;0.8] |
| Sanders et al.1 (1992) | 22:4n-6 | | 3 | 3.5(0.3) | 6 | 2.3(0.3) | 1.2 | [-1.2;0.2] |
| Sanders et al.1 (1992) | 22:5n-6 | | 3 | 0.7(0.2) | 6 | 0.5(0.3) | 0.2 | [0.5;1.9] |
| Sanders et al.1 (1992) | 22:5n-3 | | 3 | 1.7(0.4) | 6 | 1.8(0.2) | -0.1 | [-0.2;0.6] |
| Sanders et al.1 (1992) | 22:6n-3 (DHA) | | 3 | 1.9(0.3) | 6 | 6.2(0.4) | -4.3 | [-1.2;1] |
| **Cholesterol intake** | | | | | | | | |
| Hovinen et al. (2021) | cholesterol | g/d | 6 | 0.9(0.5) | 24 | 131.8(45.6) | -130.8 | [-150.1;-111.6] |
| Weder et al. (2021) | cholesterol | mg/4.184 MJ/d) | 139 | 28.4(32.8) | 164 | 99.1(25.9) | -70.7 | [-77.5;-63.9] |
| Larsson et al. 2, m (2002) | cholesterol | mg | 15 | 2.1(2.9) | 15 | 326(58) | -323.9 | [-356.1;-291.7] |
| Larsson et al. 2, f (2002) | cholesterol | mg | 15 | 2.3(2.4) | 15 | 230(76) | -227.7 | [-269.8;-185.6] |
| **Cholesterol blood** | | | | | | | | |
| Hovinen et al. (2021) | cholesterol | mmol/l | 6 | 2.8(0.2) | 24 | 4.1(0.6) | -1.3 | [-1.6;-1] |
| Desmond et al. (2021) | cholesterol | mg/dl | 52 | -32.1(46.9) | 72 | - | -32.1 | [-34.8;-32.4] |
| **Polyunsaturated fatty acids** | | | | | | | | |
| Larsson et al. 2, m (2002) | PUFA | g | 15 | 21(3.8) | 15 | 12(3.7) | 9.0 | [6.2;11.8] |
| Larsson et al. 2, f (2002) | PUFA | g | 15 | 15(6.5) | 15 | 8.6(2.9) | 6.4 | [2.6;10.2] |
| **Metabolites fatty acids** | | | | | | | | |

| | | | | | | | | |
|---|---|---|---|---|---|---|---|---|
| Hovinen et al. (2021) | carnitine 18-3 | log2 FC | 6 | 1.1(0.1) | 24 | 1(0.1) | 0.1 | [0;0.2] |
| Hovinen et al. (2021) | carnitine 18-4 | log2 FC | 6 | 1.3(0.1) | 24 | 1(0.1) | 0.3 | [0.2;0.3] |
| Hovinen et al. (2021) | carnitine 20-4 | log2 FC | 6 | 1.6(0.5) | 24 | 1(0.2) | 0.6 | [0;1.1] |
| Hovinen et al. (2021) | lysoPC 16-0 | log2 FC | 6 | 1.5(0.3) | 24 | 1(0.2) | 0.5 | [0.1;0.9] |
| Hovinen et al. (2021) | lysoPC 18-1 | log2 FC | 6 | 1.7(0.4) | 24 | 1(0.2) | 0.7 | [0.3;1.1] |
| Hovinen et al. (2021) | lysoPE 16-0 | log2 FC | 6 | 0.9(0.2) | 24 | 1(0.3) | -0.1 | [-0.3;0.2] |
| Hovinen et al. (2021) | lysoPE 18-1 | log2 FC | 6 | 1.1(0.3) | 24 | 1(0.3) | 0.1 | [-0.3;0.5] |
| Hovinen et al. (2021) | tg_c260 | log2 FC | 6 | 2.3(1.6) | 24 | 1(0.3) | 1.3 | [-0.4;3] |
| Hovinen et al. (2021) | tg_c310 | log2 FC | 6 | 2.3(1) | 24 | 1(0.5) | 1.3 | [0.2;2.4] |
| Hovinen et al. (2021) | tg_c330 | log2 FC | 6 | 2.1(0.8) | 24 | 1(0.6) | 1.1 | [0.3;2] |
| Hovinen et al. (2021) | tg_c500-o | log2 FC | 6 | 0.9(0.1) | 24 | 1(0.1) | -0.1 | [-0.2;-0.1] |
| Hovinen et al. (2021) | tg_c534 | log2 FC | 6 | 0.8(0.1) | 24 | 1(0.1) | -0.2 | [-0.3;-0.1] |
| Hovinen et al. (2021) | tg_c580-o | log2 FC | 6 | 0.7(0.1) | 24 | 1(0.3) | -0.3 | [-0.4;-0.2] |
| Hovinen et al. (2021) | tg_c584 | log2 FC | 6 | 0.9(0.1) | 24 | 1(0.1) | -0.1 | [-0.2;0] |
| Hovinen et al. (2021) | tg_c589-o | log2 FC | 6 | 1(0.1) | 24 | 1(0.1) | 0.0 | [-0.1;0.1] |
| Hovinen et al. (2021) | tg_c6210 | log2 FC | 6 | 1(0.1) | 24 | 1(0.2) | 0.0 | [-0.1;0.1] |
| **HDL** | | | | | | | | |
| Desmond et al. (2021) | HDL | % | 52 | -12.6(19.8) | 72 | - | -12.7 | [-11.2;-10] |
| Hovinen et al. (2021) | HDL | mmol/l | 6 | 1.2(0.2) | 24 | 1.4(0.2) | -0.3 | [-0.5;-0.1] |
| Alexy et al. (2021) | HDL | mg/dl | 111 | 56.1(12.9) | 136 | 57(13.1) | -0.9 | [-4.2;2.4] |
| **Triglycerides** | | | | | | | | |

| | | | | | | | | |
|---|---|---|---|---|---|---|---|---|
| Desmond et al. (2021) | triglycerides | % | 52 | 11(68.5) | 72 | - | 11 | [1.1;4.9] |
| Alexy et al. (2021) | triglycerides | mg/dl | 111 | 70(22.6) | 136 | 68.1(23.7) | 1.8 | [-4;7.7] |
| **LDL** | | | | | | | | |
| Desmond et al. (2021) | LDL | % | 52 | -20.5(40.7) | 72 | - | -20.5 | [-24.5;-22.3] |
| Hovinen et al. (2021) | LDL | mmol/l | 6 | 1.4(0.3) | 24 | 2.6(0.6) | -1.2 | [-1.6;-0.8] |
| Alexy et al. (2021) | LDL | mg/dl | 111 | 73.2(21.8) | 136 | 101.8(0) | -28.6 | [-32.7;-24.5] |
| Alexy et al. (2021) | non- HDL-cholesterol | mg/dl | 111 | 79.2(22.3) | 136 | 91.2(23.8) | -12.0 | [-17.8;-6.2] |
| Desmond et al. (2021) | VLDL | % | 52 | 6(64.9) | 72 | - | 6 | [-1.9;1.9] |
| **Cholesterol metabolites** | | | | | | | | |
| Hovinen et al. (2021) | avenasterol | µg/mg of TC | 6 | 0.7(0.1) | 24 | 0.5(0.2) | 0.2 | [0.1;0.3] |
| Hovinen et al. (2021) | campesterol | µg/mg of TC | 6 | 4.6(0.6) | 24 | 3.3(1.2) | 1.3 | [0.5;2] |
| Hovinen et al. (2021) | cholestanol | µg/mg of TC | 6 | 2(0.1) | 24 | 1.7(0.2) | 0.3 | [0.2;0.5] |
| Hovinen et al. (2021) | cholestenol | µg/mg of TC | 6 | 0.2(0) | 24 | 0.1(0) | 0.0 | [0;0.1] |
| Hovinen et al. (2021) | lathosterol | µg/mg of TC | 6 | 0.8(0.3) | 24 | 0.8(0.3) | 0.0 | [-0.3;0.3] |
| Hovinen et al. (2021) | sitosterol | µg/mg of TC | 6 | 2.6(0.5) | 24 | 1.7(0.6) | 0.9 | [0.4;1.5] |

| | | | | | | | | |
|---|---|---|---|---|---|---|---|---|
| **Bile acids** | | | | | | | | |
| Hovinen et al. (2021) | bile acid conjugation (all B | | 6 | 0.1(0.1) | 24 | 0.2(0.1) | -0.1 | [-0.2;-0.1] |
| Hovinen et al. (2021) | bile acid conjugation (prima) | | 6 | 0.1(0.1) | 24 | 0.2(0.1) | -0.1 | [-0.2;-0.1] |
| Hovinen et al. (2021) | bile acid conjugation (sec. ) | | 6 | 0(0.1) | 24 | 0.1(0.1) | -0.1 | [-0.2;0] |
| Hovinen et al. (2021) | chenodeoxycholic acid | µlmol/l | 6 | 1(1.3) | 24 | 0.1(0.1) | 0.8 | [-0.5;2.2] |
| Hovinen et al. (2021) | cholic acid | µmol/l | 6 | 0.2(0.2) | 24 | 0.1(0.1) | 0.1 | [-0.1;0.3] |
| **Vitamin A/ Retinol Equivalents / Beta Carotene intake** | | | | | | | | |
| Hovinen et al. (2021) | vitamin A | µg/d | 6 | 451.8(122.5) | 24 | 488.1(156) | -36.4 | [-170.5;97.8] |
| Weder et al. (2021) | vitamin A w/o supplements | µg/d | 139 | 597.1(329.8) | 164 | 668.3(448.6) | -71.2 | [-159.4;17.1] |
| Weder et al. (2021) | vitamin A w supplements | µg/d | 139 | 646.6(368.8) | 164 | 678.8(430.8) | -32.2 | [-122.6;58.2] |
| Larsson et al. 2, m (2002) | vitamin A | RE | 15 | 1045(273) | 15 | 1226(209) | -181.0 | [-363.5;1.5] |
| Larsson et al. 2, f (2002) | vitamin A | RE | 15 | 966(683) | 15 | 1169(588) | -203.0 | [-680.5;274.5] |

| | | | | | | | | |
|---|---|---|---|---|---|---|---|---|
| Alexy et al. (2021) | vitamin A | µg/1000kcal | 110 | 552.6(321.3) | 135 | 516.2(251.5) | 36.4 | [-37.5;110.4] |
| Weder et al. (2021) | beta-carotene w/o supplements | mg/d | 139 | 3.6(2.4) | 164 | 3.6(4) | 0.0 | [-0.7;0.7] |
| Weder et al. (2021) | beta-carotene w supplements | mg/d | 139 | 3.9(2.7) | 164 | 3.7(3.8) | 0.2 | [-0.5;1] |
| **Vitamin A blood** | | | | | | | | |
| Hovinen et al. (2021) | vitamin A | Arbitrary units | 6 | 7.8(2.9) | 24 | 18.4(6.5) | -10.6 | [-14.3;-6.8] |
| **Vitmain D intake** | | | | | | | | |
| Hovinen et al. (2021) | vitamin D | µg/d | 6 | 20.3(5.9) | 24 | 17.3(7.5) | 2.9 | [-3.5;9.4] |
| Weder et al. (2021) | vitamin D w/o supplements | µg/d | 139 | 0.7(0.4) | 164 | 1.2(1.1) | -0.5 | [-0.7;-0.3] |
| Weder et al. (2021) | Vitamin D w supplements | µg/d | 139 | 14.6(9.8) | 164 | 34.2(176.3) | -19.6 | [-46.8;7.6] |
| Larsson et al. 2, m (2002) | vitamin D | µg | 15 | 3.7(1.2) | 15 | 7.7(2.2) | -4.0 | [-5.3;-2.7] |
| Larsson et al. 2, f (2002) | vitamin D | µg | 15 | 2(1.3) | 15 | 5.1(1.6) | -3.1 | [-4.2;-2] |
| **Vitamin D blood** | | | | | | | | |
| Hovinen et al. (2021) | vitamin D | nmol/l | 6 | 47.7(7.8) | 24 | 73.5(15.8) | -25.9 | [-35.5;-16.3] |
| Hovinen et al. (2021) | vitamin D | nmol/l | 6 | 59.8(6.1) | 24 | 74.4(14.9) | -14.6 | [-22.8;-6.4] |
| Desmond et al. (2021) | vitamin D without supplements | nmol/L | 52 | -2.5(4.5) | 72 | - | -2.5 | [-3.8;-1.2] |

| | | | | | | | | |
|---|---|---|---|---|---|---|---|---|
| Desmond et al. (2021) | vitamin D3 with supplements | nmol/L | 52 | -13.2(3.4) | 72 | - | -13.2 | [-14.2;-12.2] |
| Alexy et al. (2021) | vitamin D3 with supplements | ng/ml | 111 | 26.9(10.1) | 136 | 24.9(10.1) | 1.9 | [-0.6;4.5] |
| **Zinc intake** | | | | | | | | |
| Hovinen et al. (2021) | zinc | mg/d | 6 | 10.5(3.1) | 24 | 7.8(1.7) | 2.7 | [-0.6;6.1] |
| Weder et al. (2021) | zinc w/o supplements | mg/d | 139 | 5(1.8) | 164 | 4.9(1.3) | 0.1 | [-0.2;0.5] |
| Weder et al. (2021 | zinc w supplements | mg/d | 139 | 4.9(1.9) | 164 | 4.9(1.2) | 0.0 | [-0.3;0.4] |
| Larsson et al. 2, m (2002) | zinc | mg | 15 | 10(2.4) | 15 | 16(3.3) | -6.0 | [-8.2;-3.8] |
| Larsson et al. 2, f (2002) | zinc | mg | 15 | 7.8(2.2) | 15 | 11(2.6) | -3.2 | [-5;-1.4] |
| **Zinc blood** | | | | | | | | |
| Alexy et al. (2021) | zinc | mg/1000kcal | 110 | 5.2(1.2) | 135 | 5(0.9) | 0.2 | [0;0.5] |
| Hovinen et al. (2021) | zinc | µmol/l | 6 | 14.3(4.3) | 24 | 15.3(3.6) | -0.9 | [-5.6;3.7] |
| **Biotin urine** | | | | | | | | |
| Lombard et al. 2 (1989) | urinar biotin excretion | nmol/d | 25 | 142(80) | 29 | 82(40) | 60.0 | [24.2;95.8] |
| Lombard et al. 2 (1989) | urinar biotin excretion | nmol/mmol | 25 | 36.1(18.2) | 29 | 17.2(10.3) | 18.9 | [10.6;27.2] |
| **Tocopherol/ Vit E** | | | | | | | | |

| | | | | | | | | |
|---|---|---|---|---|---|---|---|---|
| Alexy et al. (2021) | vitamin E | mg/1000kcal | 110 | 9.9(2.7) | 135 | 6.4(2.3) | 3.5 | [2.9;4.2] |
| Weder et al. (2021) | vitamin E w/o supplements | mg/d | 139 | 9.4(4.9) | 164 | 6.2(3.5) | 3.2 | [2.2;4.2] |
| Weder et al. (2021) | vitamin E w supplements | mg/d | 139 | 7.2(1.7) | 164 | 5.5(2.5) | 1.7 | [1.2;2.1] |
| Larsson et al. 2, m (2002) | vitamin E | alpha TE | 15 | 18(3.1) | 15 | 9.2(2.2) | 8.8 | [6.8;10.8] |
| Larsson et al. 2, f (2002) | vitamin E | alpha TE | 15 | 13(5.7) | 15 | 7.3(2.8) | 5.7 | [2.3;9.1] |
| **Vitamin K** | | | | | | | | |
| Weder et al. (2021) without supplement | vitamin K w/o supplements | µg/d | 139 | 83.6(20.9) | 164 | 52.5(36) | 31.1 | [24.5;37.6] |
| Weder et al. (2021) with supplement | vitamin K w supplements | µg/d | 139 | 92.7(52.8) | 164 | 50.2(32.8) | 42.5 | [32.3;52.6] |
| **Vitamin C** | | | | | | | | |
| Alexy et al. (2021) | vitamin C | mg/1000kcal | 110 | 66.9(30.4) | 135 | 53.1(35.1) | 13.8 | [5.6;22.1] |
| Weder et al. (2021) | vitamin C w/o supplements | mg/d | 139 | 64.9(29.6) | 164 | 50(26.1) | 14.9 | [8.5;21.3] |
| Weder et al. (2021) | vitamin C w supplements | mg/d | 139 | 71.9(35.9) | 164 | 49.3(23.8) | 22.5 | [15.5;29.6] |
| Larsson et al. 2, m (2002) | vitamin C | mg | 15 | 203(61) | 15 | 96(32) | 107.0 | [70;144] |
| Larsson et al. 2, f (2002) | vitamin C | mg | 15 | 178(99) | 15 | 104(44) | 74.0 | [15.5;132.5] |

| | | | | | | | | |
|---|---|---|---|---|---|---|---|---|
| **Folate / Equivalent intake** | | | | | | | | |
| Alexy et al. (2021) | folate | µg/1000kcal | 110 | 158.9(45.7) | 135 | 104.3(32) | 54.7 | [44.5;64.8] |
| Hovinen et al. (2021) | folate | µg/d | 6 | 490.8(58.3) | 24 | 164(50.6) | 326.9 | [265.5;388.2] |
| Weder et al. (2021) | folate w/o supplements | µg/d | 139 | 159.4(77.7) | 164 | 115.2(34.6) | 44.1 | [30.1;58.2] |
| Weder et al. (2021) | folate w supplements | µg/d | 139 | 170.8(93.4) | 164 | 115.7(37.2) | 55.1 | [38.5;71.8] |
| Larsson et al. 2, m (2002) | folate | µg | 15 | 551(142) | 15 | 263(42) | 288.0 | [206.9;369.1] |
| Larsson et al. 2, f (2002) | folate | µg | 15 | 473(187) | 15 | 226(73) | 247.0 | [138.1;355.9] |
| **Folate blood** | | | | | | | | |
| Svetnicka et al. (2022) | folate | µg/L | 69 | 17.4(4.6) | 52 | 16.1(7.7) | 1.3 | [-1.1;3.7] |
| Alexy et al. (2021) | folate | µg/L | 111 | 326.1(58.1) | 136 | 292.1(56.8) | 33.9 | [19.4;48.4] |
| Hovinen et al. (2021) | folate | nmol/l | 6 | 1036.2(157) | 24 | 606.1(195.3) | 430.1 | [259.3;600.8] |
| **Vitamin B1/ Thiamine** | | | | | | | | |
| Alexy et al. (2021) | vitamin B1 | µg/1000kcal | 110 | 594.5(155.3) | 135 | 487.6(112.2) | 106.9 | [72.1;141.8] |
| Weder et al. (2021) | vitamin B1 w/o supplements | µg/d | 139 | 630.1(273.1) | 164 | 523.8(166.4) | 106.3 | [53.9;158.6] |
| Weder et al. (2021) | vitamin B1 w supplements | µg/d | 139 | 645.5(271.2) | 164 | 518.4(166.9) | 127.1 | [75;179.2] |

| | | | | | | | | |
|---|---|---|---|---|---|---|---|---|
| Larsson et al. 2, m (2002) | vitamin B1 | mg | 15 | 1.9(0.6) | 15 | 2.2(0.5) | -0.3 | [-0.7;0.1] |
| Larsson et al. 2, f (2002) | vitamin B1 | mg | 15 | 1.5(0.8) | 15 | 1.5(0.5) | 0.0 | [-0.5;0.5] |
| **Vitamin B2/ Riboflavin intake** | | | | | | | | |
| Alexy et al. (2021) | vitmain B2 | µg/1000kcal | 110 | 559.6(155.3) | 135 | 487.6(112.2) | 72 | [-583.9;-535.3] |
| Weder et al. (2021) | vitamin B2 w/o supplements | µg/d | 139 | 432.8(157.5) | 164 | 681.5(221.5) | -248.6 | [-291.6;-205.6] |
| Weder et al. (2021) | vitamin B2 w supplements | µg/d | 139 | 473.2(204) | 164 | 688.3(238.6) | -215.1 | [-265.2;-165.1] |
| Larsson et al. 2, m (2002) | vitmain B2 | mg | 15 | 1.2(0.6) | 15 | 2.8(0.7) | -1.6 | [-2.1;-1.1] |
| Larsson et al. 2, f (2002) | vitmain B2 | mg | 15 | 1.1(0.5) | 15 | 1.9(0.7) | -0.8 | [-1.3;-0.3] |
| **Vitamin B2 blood** | | | | | | | | |
| Alexy et al. (2021) | vitamin B2 | µg/l | 111 | 190(34.9) | 136 | 207.3(25.4) | -17.3 | [-25.1;-9.5] |
| **Vitamin B6** | | | | | | | | |
| Weder et al. (2021) | vitamin B6 w/o supplements | mg/d | 139 | 0.9(0.4) | 164 | 0.8(0.2) | 0.1 | [0.1;0.2] |
| Weder et al. (2021) | vitamin B6 w supplements | mg/d | 139 | 0.9(0.4) | 164 | 0.8(0.2) | 0.1 | [0.1;0.2] |
| Larsson et al. 2, m (2002) | vitamin B6 | mg | 15 | 2.7(0.5) | 15 | 2.8(0.6) | -0.1 | [-0.5;0.3] |
| Larsson et al. 2, f (2002) | vitamin B6 | mg | 15 | 2.1(0.7) | 15 | 2(0.6) | 0.1 | [-0.4;0.6] |
| **Vitamin B12 intake** | | | | | | | | |

| | | | | | | | | |
|---|---|---|---|---|---|---|---|---|
| Alexy et al. (2021) | vitamin B12 | µg/1000kcal | 110 | NA | 135 | 1.6(0.6) | | |
| Weder et al. (2021) | vitamin B12 w/o supplements | µg/d | 139 | 0.3(0.6) | 164 | 1.8(1.1) | -1.5 | [-1.7;-1.3] |
| Weder et al. (2021) | vitamin B12 w supplements | µg/d | 139 | 181.9(246) | 164 | 2(1.5) | 179.8 | [138.6;221.1] |
| Larsson et al. 2, m (2002) | vitamin B12 | µg | 15 | 0.1(0) | 15 | 5.9(1.5) | -5.8 | [-6.6;-5] |
| Larsson et al. 2, f (2002) | vitamin B12 | µg | 15 | 0(0.1) | 15 | 5(2.5) | -5.0 | [-6.4;-3.6] |
| **Vitamin B12 blood** | | | | | | | | |
| Svetnicka et al. (2022) | vitamin B12 a | pmol/l | 69 | 118.6(64.1) | 52 | 104.2(53.7) | 14.4 | [-6.9;35.6] |
| Svetnicka et al. (2022) | vitamin B12 | µg/l | 69 | 622.5(324.8) | 52 | 482.3(170.9) | 140.2 | [49.5;230.8] |
| Desmond et al. (2021) | vitamin B12 supplement | pmol/l | 52 | -104(43.8) | 72 | - | -104.0 | [52.6;81.2] |
| Desmond et al. (2021) | vitamin B12 no supplement | pmol/l | 52 | -183.8(33.9) | 72 | - | -217.6 | [-116.2;-91.8] |
| **Homocystein** | | | | | | | | |
| Desmond et al. (2021) | homocystein w/o supplements | % | 52 | 48(12) | 72 | - | 48.0 | [-11.8;-8.2] |
| Desmond et al. (2021) | Homocystein w supplements | % | 52 | 14(11) | 72 | - | 14.0 | [44.7;51.3] |
| Alexy et al. (2021) | MMA | nmol/l | 111 | 143.6(51.1) | 136 | 161.4(61.5) | -17.7 | [-31.8;-3.6] |
| **Holo Transcobalamin** | | | | | | | | |

| | | | | | | | | |
|---|---|---|---|---|---|---|---|---|
| Alexy et al. (2021) | holo transcobalamin | pmol/l | 111 | 92.2(70.2) | 136 | 69.1(27.5) | 23.1 | [9.1;37.1] |
| **Calcium** | | | | | | | | |
| Alexy et al. (2021) | calcium | mg/1000kcal | 110 | 346.4(155.8) | 135 | 406.5(109.4) | -60.1 | [-94.8;-25.4] |
| Weder et al. (2021) | calcium w/o supplements | mg/d | 139 | 366.2(174.2) | 164 | 464.4(162.3) | -98.2 | [-136.5;-59.9] |
| Weder et al. (2021) | calcium w supplements | mg/d | 139 | 363.7(167.1) | 164 | 467.1(165.1) | -103.4 | [-141.1;-65.6] |
| Larsson et al. 2, m (2002) | calcium | mg | 15 | 517(158) | 15 | 1697(444) | -1180.0 | [-1436.7;-923.3] |
| Larsson et al. 2, f (2002) | calcium | mg | 15 | 538(350) | 15 | 1328(372) | -790.0 | [-1060.6;-519.4] |
| **Bone** | | | | | | | | |
| Desmond et al. (2021) | total body Bone mineral content ( TBLH BMC) | change | 52 | -3.7(12) | 72 | - | -3.7 | [-17.5;-15.3] |
| Desmond et al. (2021) | L2-L4 BMC | change | 52 | -5.6(18) | 72 | - | -5.6 | [-12.5;-8.5] |
| Desmond et al. (2021) | bone apparent mineral density (BMAD) z-Score | change | 52 | -0.6(1.7) | 72 | - | -0.6 | [-0.7;-0.6] |
| **Phosphorous** | | | | | | | | |
| Larsson et al. 2, m (2002) | phosphorous | mg | 15 | 1361(280) | 15 | 2176(408) | -815.0 | [-1078.7;-551.3] |
| Larsson et al. 2, f (2002) | phosphorous | mg | 15 | 1025(304) | 15 | 1536(378) | -511.0 | [-768.4;-253.6] |

| | | | | | | | | |
|---|---|---|---|---|---|---|---|---|
| **Magnesium** | | | | | | | | |
| Alexy et al. (2021) | magnesium | mg/1000kcal | 110 | 261.6(80) | 135 | 159.2(33.1) | 102.4 | [86.3;118.5] |
| Weder et al. (2021) | magnesium w/o supplements | mg/d | 139 | 251.7(98.7) | 164 | 165.5(45.9) | 86.2 | [68.2;104.2] |
| Weder et al. (2021) | magnesium w supplements | mg/d | 139 | 247.7(96.3) | 164 | 154.9(36) | 92.8 | [75.8;109.9] |
| Larsson et al. 2, m (2002) | magnesium | mg | 15 | 559(96) | 15 | 467(105) | 92.0 | [16.6;167.4] |
| Larsson et al. 2, f (2002) | magnesium | mg | 15 | 443(120) | 15 | 325(77) | 118.0 | [41.8;194.2] |
| **Iron intake** | | | | | | | 0.0 | |
| Alexy et al. (2021) | iron | mg/1000kcal | 110 | 9.3(2.3) | 135 | 5.9(1.1) | 3.4 | [2.9;3.8] |
| Hovinen et al. (2021) | iron | mg/d | 6 | 12.2(1.7) | 24 | 7.4(1.9) | 4.8 | [3;6.7] |
| Weder et al. (2021) | iron w/o supplements | mg/d | 139 | 8.7(3.4) | 164 | 6.1(1.9) | 2.6 | [2;3.2] |
| Weder et al. (2021) | iron w supplements | mg/d | 139 | 8.8(3.4) | 164 | 6.1(2) | 2.7 | [2;3.2] |
| Larsson et al. 2, m (2002) | iron | mg | 15 | 18(31) | 15 | 18(4.8) | 0.0 | [-17.4;17.4] |
| Larsson et al. 2, f (2002) | iron | mg | 15 | 14(4.5) | 15 | 11(3) | 3.0 | [0.1;5.9] |
| **Ferritin** | | | | | | | | |
| Desmond et al. (2021) | ferritin | % | 52 | -25(68.5) | 72 | - | -25 | [-30.9;-25.1] |
| Alexy et al. (2021) | ferritin | µg/l | 111 | 68.5(16.7) | 136 | 39.6(19.3) | 28.9 | [-11;-2] |

| | | | | | | | | |
|---|---|---|---|---|---|---|---|---|
| Hovinen et al. (2021) | ferritin | µg/l | 6 | 20.2(14.5) | 24 | 17.9(9.3) | 2.3 | [-12.9;17.5] |
| **Potassium** | | | | | | | | |
| Weder et al. (2021) | potassium w/o supplements | mg/d | 139 | 1759.9(523.7) | 164 | 1602.4(443) | 157.5 | [46.6;268.3] |
| Weder et al. (2021) | potassium w supplements | mg/d | 139 | 1763.4(512.6) | 164 | 1622.8(420.5) | 140.6 | [33.4;247.9] |
| Larsson et al. 2, m (2002) | potassium | mg | 15 | 4200(863) | 15 | 4100(694) | 100.0 | [-487.8;687.8] |
| Larsson et al. 2, f (2002) | potassium | mg | 15 | 3460(1240) | 15 | 3160(921) | 300.0 | [-521.4;1121.4] |
| **Iodine intake** | | | | | | | | |
| Weder et al. (2021) | iodine w/o supplements | µg/d | 139 | 37.2(22.5) | 164 | 50.5(20.2) | -13.3 | [-18.2;-8.4] |
| Weder et al. (2021) | iodine w supplements | µg/d | 139 | 38.3(20.1) | 164 | 51.8(21) | -13.5 | [-18.2;-8.9] |
| **Iodine** | | | | | | | | |
| Hovinen et al. (2021) | iodine to creatinine (urine) | µg/l per mmol/l | 6 | 41.5(32.7) | 24 | 48.6(22.7) | -7.1 | [-41.7;27.5] |
| **Niacin Vitamin B3** | | | | | | | | |
| Larsson et al. 2, m (2002) | niacin | NE | 15 | 34(7.8) | 15 | 48(9.5) | -14.0 | [-20.5;-7.5] |
| Larsson et al. 2, f (2002) | niacin | NE | 15 | 26(8.4) | 15 | 32(8) | -6.0 | [-12.1;0.1] |
| **Sodium** | | | | | | | | |
| Larsson et al. 2, m (2002) | sodium | mg | 15 | 3797(856) | 15 | 4656(1108) | -859.0 | [-1602.1;-115.9] |
| Larsson et al. 2, m (2002) | sodium | mg | 15 | 2580(986) | 15 | 3040(614) | -460.0 | [-1080.4;160.4] |

| | | | | | | | | |
|---|---|---|---|---|---|---|---|---|
| **Selenium** | | | | | | | | |
| Larsson et al. 2, m (2002) | selenium | µg | 15 | 10(3) | 15 | 27(7.6) | -17.0 | [-21.4;-12.6] |
| Weder et al. (2023) | selenium | µg | 139 | 18.8(8.0) | 164 | 20.9(5.3) | -2.1 | [-3.7; -0.6] |
| **Albumin** | | | | | | | | |
| Lombard et al. 1 (1989) | albumin serum | g/l | 25 | 45.6(3.6) | 29 | 72.9(2.5) | -27.3 | [-29;-25.6] |
| **MCV** | | | | | | | | |
| Svetnicka et al. (2022) | MCV | fl | 69 | 79.9(0) | 52 | 80.2(0) | -0.3 | |
| Desmond et al. (2021) | MCV<br>w supplements | fl | 52 | 0.8(1.2) | 72 | - | 0.8 | [0.7;1.1] |
| Desmond et al. (2021) | MCV<br>w/osupplements | fl | 52 | 4.3(1.4) | 72 | - | 4.3 | [0.5;1.2] |
| **Hemoglobine** | | | | | | | | |
| Alexy et al. (2021) | HGB | g(dl | 111 | 13.4(1) | 136 | 13.5(1) | -0.1 | [-0.4;0.2] |
| Desmond et al. (2021) | HGB | g/dl | 52 | -0.4(1.2) | 72 | - | -0.4 | [-0.4;-0.3] |
| **Others** | | | | | | | | |
| Hovinen et al. (2021) | transthyretin | mg/l | 6 | 146.2(13.7) | 24 | 183.4(23.1) | -37.2 | [-53;-21.4] |
| Hovinen et al. (2021) | RBP | RE(µmol/l) | 6 | 0.9(0.1) | 24 | 1.3(0.2) | -0.4 | [-0.5;-0.2] |
| Hovinen et al. (2021) | TfR | mg/l | 6 | 6.5(0.5) | 24 | 6.6(2) | -0.1 | [-1.1;0.8] |
| Desmond et al. (2021) | RBC Erythrocytes | M/µl | 52 | -0.2(0.4) | 72 | - | -0.2 | [-0.2;-0.2] |
| **Carnitin** | | | | | | | | |

| | | | | | | | | |
|---|---|---|---|---|---|---|---|---|
| Lombard et al. 1 (1989) | plasma carnitin free | µgmol/l | 13 | 26.1(7.6) | 15 | 38.7(7.5) | -12.6 | [-18.5;-6.7] |
| Lombard et al. 1 (1989) | plasma carnitin free | µgmol/l | 12 | 25.8(5.2) | 14 | 34.4(6.3) | -8.6 | [-13.3;-3.9] |
| Lombard et al. 1 (1989) | plasma carnitin free | µgmol/l | 13 | 32(8.5) | 15 | 46.8(9.1) | -14.8 | [-21.7;-7.9] |
| Lombard et al. 1 (1989) | plasma carnitin free | µmol/l | 12 | 33.9(5.8) | 14 | 45.9(5.6) | -12.0 | [-16.6;-7.4] |
| Lombard et al. 1 (1989) | urinary carnitin free | µmol*kg-1xd-1 | 13 | 0.1(0.1) | 15 | 4.1(2.6) | -3.9 | [-5.4;-2.5] |
| Lombard et al. 1 (1989) | urinary carnitin total | µmolxkg-1xd-1 | 13 | 1.2(0.3) | 15 | 8.6(4.8) | -7.4 | [-10;-4.8] |
| Lombard et al. 1 (1989) | urinary carnitin free | µmolxkg-1xd-1 | 12 | 0.1(0.1) | 14 | 2.5(1.5) | -2.4 | [-3.2;-1.6] |
| Lombard et al. 1 (1989) | urinary carnitin total | µmolxkg-1xd-1 | 12 | 1.2(0.5) | 14 | 5.7(2.2) | -4.5 | [-5.8;-3.2] |
| **Inflammation** | | | | | | | | |
| Desmond | Molar IGF-1/ IGFBP-3 Ratio | | 52 | 0(0.1) | 72 | - | 0.0 | |
| Desmond | cIMT | mm | 52 | 0(0.1) | 72 | - | 0.0 | |
| **CRP** | | | | | | | | |
| Desmond et al. (2021) | hs CRP | % | 52 | -44.9(132.7) | 72 | - | -44.9 | [-51.4;-42.6] |
| **HTC** | | | | | | | | |
| Desmond et al. (2021) | HTC | % | 52 | -105(356.9) | 72 | - | -105.0 | [-118.5;-91.5] |
| **IGFs** | | | | | | | | |
| Desmond et al. (2021) | IGF-1 | ng/ml | 52 | 20(122.6) | 72 | - | 20.0 | [-18.3;-9.7] |

| | | | | | | | | |
|---|---|---|---|---|---|---|---|---|
| Desmond et al. (2021) | IGFBP-3 | ng/ml | 52 | -50(962.7) | 72 | - | -50.0 | [-138.9;-71.1] |
| **Glucose** | | | | | | | | |
| Desmond et al. (2021) | glucose | mg/dl | 52 | 2.7(10.8) | 72 | - | 2.7 | [-32.9;-28.5] |
| Desmond et al. (2021) | HOMA-IR | % | 52 | 14.9(53.4) | 72 | - | 14.9 | [2.9;6.5] |
| **Insulin** | | | | | | | | |
| Desmond et al. (2021) | insulin | µlU/ml | 52 | 0.7(3.6) | 72 | - | 0.7 | [-0.2;0.1] |
| **Anthropometry** | | | | | | | | |
| **Height** | | | | | | | | |
| Wirnitzer et al. (2021 | height | cm | 633 | 162(12.4) | 7421 | 167.1(10.4) | -5.1 | [-6.1;-4.1] |
| Alexy et al. (2021) | height | cm | 114 | 152(19) | 137 | 156(20) | -4.0 | [-8.9;0.9] |
| Hovinen et al. (2021) | height | z-score | 6 | | 24 | | | |
| Hovinen et al. (2021) | height | cm | 6 | 102.9(14) | 24 | 102.4(13.7) | 0.5 | [-14.5;15.5] |
| Larsson et al. 1, m (2002) | height | cm | 9 | 180(7) | 9 | 180(6) | 0.0 | [-6.6;6.6] |
| Larsson et al. 1, f (2002) | height | cm | 7 | 167(5) | 7 | 168(4) | -1.0 | [-6.3;4.3] |
| Weder et al. (2021) | height for age | z-score | 139 | 0(1.3) | 164 | 0.1(1) | -0.1 | [-0.4;0.1] |
| Weder et al. (2021) | height | cm | 139 | 85.6(8.8) | 164 | 88.2(9.3) | -2.6 | [-4.6;-0.6] |
| Svetnicka et al. (2022) | height | percentile | 69 | 53.1(33) | 52 | 47(26.9) | 6.0 | [-4.8;16.8] |
| Desmond et al. (2021) | height | z-score | 52 | -0.6(0.2) | 72 | - | -0.6 | [-0.6;-0.5] |
| Ferrara et al. (2021) | birth length | cm | 21 | 50.7(1.7) | 21 | 49.7(2.4) | 1.0 | [-2,3;0,3] |
| Ferrara et al. (2021) | percentile birth length | percentile | 21 | 68.5(26.1) | 21 | 50.3(34.9) | 18.2 | [-37,5;1] |

| | | | | | | | | |
|---|---|---|---|---|---|---|---|---|
| Ferrara et al. (2021) | height 6 months | cm | 21 | 66.9(4) | 21 | 67(2.8) | 0.0 | [-2,1;2,2] |
| Ferrara et al. (2021) | height 6 months | percentile | 21 | 54.6(30.7) | 21 | 47.5(30.8) | 7.0 | [-26,3;12,2] |
| Ferrara et al. (2021) | height 12 months | cm | 21 | 77.1(4.4) | 21 | 74.9(4) | 2.2 | [-4,9;0,4] |
| Ferrara et al. (2021) | height 12 months | percentile | 21 | 66.5(27.2) | 21 | 43.2(33.2) | 23.2 | [-42,2;-4,3] |
| **Weight** | | | | | | | | |
| Svetnicka et al. (2022) | weight | percentile | 69 | 40(19.2) | 52 | 41.9(17.7) | -1.8 | [-8.5;4.8] |
| Wirnitzer et al. (2021 | weight | kg | 633 | 53.5(14.8) | 7421 | 59.1(14.4) | -5.6 | [-6.8;-4.4] |
| Alexy et al. (2021) | weight | kg | 114 | 43(16) | 137 | 46(17) | -3.0 | [-7.1;1.1] |
| Hovinen et al. (2021) | Weight | | 6 | 16.2(1.8) | 24 | 16.3(1.7) | -0.1 | [-2;1.8] |
| Larsson et al. 1, m (2002) | weight | kg | 9 | 66.1(6.2) | 9 | 70.2(4.7) | -4.1 | [-9.7;1.5] |
| Larsson et al. 1, f (2002) | weight | kg | 7 | 70.7(8.8) | 7 | 58.7(7.7) | 12.0 | [2.3;21.7] |
| Weder et al. (2021) | weight | kg | 139 | 12(2.5) | 164 | 12.7(2.6) | -0.7 | [-1.3;-0.1] |
| Weder et al. (2021) | weight-for-height | z-score | 139 | 0.2(1.1) | 164 | 0.2(1) | -0.1 | [-0.3;0.2] |
| Weder et al. (2021) | weight for age | z-score | 139 | 0.1(0.9) | 164 | 0.3(0.9) | -0.1 | [-0.3;0.1] |
| Ferrara et al. (2021) | weight at 6 months | g | 21 | 7948.6(660.8) | 21 | 7265.7(929.9) | 682.9 | [-1187,7;-178] |
| Ferrara et al. (2021) | weight 12 months | percentile | 21 | 60.9(24.2) | 21 | 35.2(29.7) | 25.7 | [-42,6;-8,8] |
| Ferrara et al. (2021) | weight at 12 months | g | 21 | 10570(1344.2) | 21 | 9252.4(1042.9) | 1317.6 | [-2069,9;-565,4] |
| Ferrara et al. (2021) | weight 6 months | percentile | 21 | 77.6(19.1) | 21 | 45.7(29.6) | 31.9 | [-47,5;-16,3] |
| **Head circumference** | | | | | | | | |

| | | | | | | | | |
|---|---|---|---|---|---|---|---|---|
| Ferrara et al. (2021) | head circumference birth | cm | 21 | 34.8(2) | 21 | 34.1(1.5) | 0.7 | [-1,8;0,3] |
| Ferrara et al. (2021) | head circumference birth | percentile | 21 | 57.5(28.1) | 21 | 47.6(31.9) | 9.9 | [-28,6;8,9] |
| Ferrara et al. (2021) | head circumference 6 months | cm | 21 | 42.5(1.5) | 21 | 43(1.7) | -0.5 | [-0,5;1,5] |
| Ferrara et al. (2021) | head circumference 6 months | percentile | 21 | 46.2(28.3) | 21 | 58.1(28) | -11.9 | [-5,7;29,4] |
| Ferrara et al. (2021) | head circumference 12 months | cm | 21 | 46(1.4) | 21 | 45.8(1.7) | 0.2 | [-1,1;0,8] |
| Ferrara et al. (2021) | head circumference 12 months | percentile | 21 | 65.2(26.9) | 21 | 60.3(26.3) | 4.9 | [-21,5;11,7] |
| **Waist** | | | | | | | | |
| Desmond et al. (2021) | waist girth | z-score | 52 | -0.3(1.3) | 72 | - | -0.3 | [-0.3;-0.2] |
| **BMI** | | | | | | | | |
| Alexy et al. (2021) | BMI SDS | BMI SDS | 114 | -0.6(0.9) | 137 | | | |
| Hovinen et al. (2021) | BMI | | 6 | | 24 | | | |
| Larsson et al. 1, m (2002) | BMI | kg/m2 | 9 | 20.5(1.7) | 9 | 21.6(1) | -1.1 | [-2.5;0.3] |
| Larsson et al. 1, f (2002) | BMI | kg/m2 | 7 | 25.4(3.8) | 7 | 20.9(2.5) | 4.5 | [0.7;8.3] |
| Desmond et al. (2021) | BMI | z-score | 52 | -0.5(0.2) | 72 | - | -0.5 | [-0.5;-0.5] |
| **Skinfold** | | | | | | | | |

| | | | | | | | | |
|---|---|---|---|---|---|---|---|---|
| Desmond et al. (2021) | subscapular skinfold | z-score | 52 | -0.2(1.6) | 72 | - | -0.2 | [-4.6;3.9] |
| Desmond et al. (2021) | suprailiac skinfold | z-score | 52 | -0.6(1.4) | 72 | - | -0.6 | [-0.5;-0.4] |
| Desmond et al. (2021) | thigh girth | z-score | 52 | -0.6(1.4) | 72 | - | -0.6 | [-0.7;-0.6] |
| Desmond et al. (2021) | triceps skinfold | z-score | 52 | -0.5(1.4) | 72 | - | -0.5 | [-0.6;-0.5] |
| Desmond et al. (2021) | biceps skinfold | z-score | 52 | -0.2(1.4) | 72 | - | -0.2 | [-0.3;-0.2] |
| **Others** | | | | | | | | |
| Desmond et al. (2021) | fat mass index | | 52 | -0.7(1.4) | 72 | - | -0.7 | [-0.8;-0.7] |
| Larsson et al. 1, m (2002) | body water by doubly labeled water | kg | 9 | 39.8(3.7) | 9 | 43.3(1.4) | -3.5 | [-6.4;-0.6] |
| **Stunning/ Anemia** | | | | Mean(95%CI) | | | | |
| Headey et al. (2020) | proportion stunted | | NA | 0.455(0.432, 0.478) | NFHS Survey data | - | - | |
| Headey et al. (2020) | proportion wasted | | NA | 0.215(0.196, 0.234) | NFHS Survey data | - | - | |
| Headey et al. (2020) | proportion anemic | | NA | 0.638(0.615, 0.662) | NFHS Survey data | - | - | |
| **Others** | | | | | | | | |

| | | | | | | | | |
|---|---|---|---|---|---|---|---|---|
| Desmond et al. (2021) | gestation age | week | 52 | 38.8(1.9) | 72 | 39(1.5) | -0.2 | |
| Larsson et al. 1, m (2002) | BMR (basal metabolic range) | MJ | 9 | 7.4(0.4) | 9 | 7.7(0.5) | -0.3 | [-0.7;0.2] |
| Larsson et al. 1, m (2002) | FIL (food intake level (reported energy intake /BMR) | | 9 | 1.6(0.2) | 9 | 1.7(0.3) | -0.1 | [-0.4;0.1] |
| Larsson et al. 1, m (2002) | PAL (reported) (Physical activitiy leven (energy expenditure/BMR) | | 9 | 1.6(0.2) | 9 | 1.7(0.2) | -0.1 | [-0.2;0.1] |
| Larsson et al. 1, m (2002) | PAL (measured) | | 9 | 1.9(0.4) | 9 | 2.1(0.3) | -0.2 | [-0.5;0.2] |
| Larsson et al. 1, m (2002) | FIL/PAL(m) | | 9 | 0.9(0.2) | 9 | 0.9(0.2) | 0.0 | [-0.2;0.2] |
| Larsson et al. 1, m (2002) | PAL®/PAL(m) | | 9 | 0.9(0.2) | 9 | 0.8(0.1) | 0.1 | [-0.1;0.2] |
| Larsson et al. 1, m (2002) | Nredx0.81)/Nmeas (N=Nitrogen) | | 9 | 1(0.2) | 9 | 1(0.1) | 0.0 | [-0.2;0.2] |
| Larsson et al. 1, f (2002) | BMR (basal metabolic range) | | 7 | 6.8(0.6) | 7 | 6.1(0.5) | 0.7 | [0;1.3] |
| Larsson et al. 1, f (2002) | FIL (food intake level (reported energy intake /BMR) | | 7 | 1.2(0.3) | 7 | 1.7(0) | -0.5 | [-0.8;-0.2] |

| | | | | | | | | |
|---|---|---|---|---|---|---|---|---|
| Larsson et al. 1, f (2002) | PAL (reported) (Physical activitiy leven (energy expenditure/BMR) | | 7 | 1.5(0.1) | 7 | 1.7(0.1) | -0.2 | [-0.3;-0.1] |
| Larsson et al. 1, f (2002) | PAL (measured) | | 7 | 1.4(0.2) | 7 | 1.8(0.4) | -0.4 | [-0.9;0] |
| Larsson et al. 1, f (2002) | FIL/PAL(m) | | 7 | 0.8(0.3) | 7 | 0.9(0.2) | -0.1 | [-0.3;0.2] |
| Larsson et al. 1, f (2002) | PAL®/PAL(m) | | 7 | 1.1(0.2) | 7 | 0.9(0.2) | 0.2 | [-0.1;0.4] |
| Larsson et al. 1, f (2002) | Nredx0.81)/Nmeas (N=Nitrogen) | | 7 | 0.8(0.2) | 7 | 1(0.1) | -0.2 | [-0.4;-0.1] |
| Larsson et al. 2, m (2002 | EI(rep)/EE(mea) | | 15 | 0.9(0.2) | 14 | 0.9(0.2) | 0.0 | [-0.1;0.2] |
| Larsson et al. 2, m (2003 | (N(rep)x0.81/N(me n) | | 15 | 1.1(0.3) | 14 | 1(0.1) | 0.1 | [-0.1;0.3] |
| Larsson et al. 2, m (2004 | Na(rep)/Na(mea) | | 15 | 1(0.3) | 14 | 1.2(0.3) | -0.1 | [-0.4;0.1] |
| Larsson et al. 2, m (2005 | K(rep)=..73 or =.77)/K(mea) | | 15 | 1(0.3) | 14 | 0.9(0.2) | 0.1 | [-0.1;0.3] |
| Larsson et al. 1, f (2002 | EI(rep)/EE(mea) (energy intake/energy expenditure) | | 15 | 0.8(0.3) | 15 | 0.9(0.2) | -0.1 | [-0.2;0.1] |
| Larsson et al. 1, f (2003 | (N(rep)x0.81/N(me n) | | 15 | 1(0.3) | 15 | 1(0.1) | 0.0 | [-0.2;0.2] |
| Larsson et al. 1, f (2004 | Na(rep)/Na(mea) | | 15 | 0.8(0.4) | 15 | 1(0.2) | -0.2 | [-0.5;0] |

| | | | | | | | | |
|---|---|---|---|---|---|---|---|---|
| Larsson et al. 1, f (2005 | K(rep)=..73 or =.77)/K(mea) | | 15 | 0.9(0.3) | 15 | 0.9(0.2) | 0.0 | [-0.2;0.2] |
| Wirnitzer et al. (2021 | club sport | prevalence | 633 | 43.5(0) | 7421 | 42.5(0) | 1.0 | |
| Wirnitzer et al. (2021 | fluid intake | prevalence | 633 | 24.2(0) | 7421 | 23(0) | 1.2 | |
| Wirnitzer et al. (2021 | leisure time | prevalence | 633 | 86.4(0) | 7421 | 82(0) | 4.4 | |
| **RCT** | | | | | | | | |
| Macknin et al. (2021) W52 | weight | kg | 25 | 5.6(6.3) | 27 | | NS | |
| Macknin et al. (2021) W52 | waist | cm | 25 | -8.2(7.3) | 27 | -8.9(4.3) | 0.01 | |
| Macknin et al. (2021) W52 | syst BP | mmHg | 25 | -20.7(17.3) | 27 | -7.3(6.3) | 0.01 | |
| Macknin et al. (2021) W52 | diast BP | mmHg | 25 | -8.7(9.4) | 27 | -9.3(11) | 0.02 | |
| Macknin et al. (2021) W52 | cholesterol | mg/dL(change) | 25 | -27.3(33) | 27 | -17.7(9.4) | 0.00 | |
| Macknin et al. (2021) W52 | LDL | mg/dl | 25 | -21(14.2) | 27 | -13.7(7.8) | 0.00 | |

| | | | | | | | |
|---|---|---|---|---|---|---|---|
| Macknin et al. (2021) W52 | glucose | mg/dl | 25 | -9.3(10.2) | 27 | -9(10.2) | 0.02 |
| Macknin et al. (2021) W52 | MPO | pmol/l | 25 | -113(62.9) | 27 | -80.3(36) | 0.04 |

Abbrevations: kcal: kilocalorie; kJ: kilojoule; MJ: megajoule; g: gramm; mg: miligramm; µg: mikrogramm; mmol: milimol; µmol: mikromol; nmol: nanomol; pmol: picomol; TC: total cholesterol; fl: femtoliter; cm: centimeter; kg: kilogramm; mmHg: milimeter mercury; NA: not applicable

Desmond et al. only provided differences between the two groups, therefore there are no values in columns vgn/omn.

Headey et al. did not provide the number of vegans/ omnivores, therefore Means(95%CIs) are taken from the original publication.

In the Study from Macknin et al. 2021 all values given in the results are change scores after a 52 week intervention.

Table S7: Results of the risk of bias assessment using ROBIN-E and RoB 2.0

ROBINS-E

| **Study** | **Confounding** | **Measurement of exposure** | **Selection of participantnts** | **Post-exposure elaborations** | **Missing data** | **Measurment Of Outcomes** | **Selection Bias** | **Overall risk** | **Explanation** |
|---|---|---|---|---|---|---|---|---|---|
| *Svetnicka et al. 2022* | | | | | | | | | Only adjusted for sex and age |
| *Wirnitzer et al. 2021* | | | | | | | | | Only questionnaire, not many confounders |
| *Desmond et al. 2021* | | | | | | | | | - |
| *Alexy et al 2021* | | | | | | | | | Some concerns for nutrient intakes -□ used questionnaires |
| *Hovinen et al. 2021* | | | | | | | | | Only sex/age, some dropouts |
| *Weder et al. 2021* | | | | | | | | | Questionnaire food intake |
| *Weder et al. 2023* | | | | | | | | | Food protocol |

| | | | | | | | | | |
|---|---|---|---|---|---|---|---|---|---|
| *Weder et al. 2019* | | | | | | | | | See Alexy |
| *Pawlak et al. 2014* | | | | | | | | | No adjustments for confounders |
| *Larsson et al. -1, 2002* | | | | | | | | | Only age sex height |
| *Larsson et al.-2, 2002* | | | | | | | | | Only age sex height |
| *Sanders et al. 1992* | | | | | | | | | No adjustments for confounders |
| *Headey et al. 2020* | | | | | | | | | Only assessed nutrition of mothers |
| *Lombard et al. -1, 1989* | | | | | | | | | No adjustments for confounders |
| *Lombard et al. – 2, 1989* | | | | | | | | | Only adjusted for sex, Adventists, questionnaire |
| *Ferrara et al. 2021* | | | | | | | | | No adjustments for confounders |

RoB 2.0

| **Study** | **Randomization process** | **Assignement to intervention** | **Adherence to intervention** | **Missing data** | **Measurement of outcome** | **selectionbias** | **Overall risk** | **Explenation** |
|---|---|---|---|---|---|---|---|---|
| *Macknin et al. 2021* | | | | | | | | Bad adherence, high dropout rate |

| | | | | |
|---|---|---|---|---|
| low | some | high | Very high | Not assessed |

**Supplementary information about RoB 2.0.**

RoB-2.0 (risk of bias tool for randomized trials) contains the following five domains. (1.) Risk of bias arising from randomization process (2.) Risk of bias due to deviations from the intended interventions (effect of assignment to intervention and effect of adhering to intervention) (3.) Missing outcome data (4.) Risk of bias in measurement of the outcome (5.) Risk of bias in selection of the reported results

Signaling questions which must be answered with yes (Y), probably yes (PY), probably no (PN), no (N) or no information (NI) help to access the risk of bias of each domain. The overall risk of bias was evaluated by the researchers (AK, JK) by evaluating the results of the five domains.

**Supplementary information about ROBINS-E**

The following seven Domains are part of ROBINS-E (1.) Risk of bias due to confounding, (2.) Risk of bias arising from measurement of the exposure (3.) Risk of bias in selection of participants into the study (or into the analysis) (4.) Risk of bias due to post-exposure interventions (5.) Risk of bias due to missing data (6.) Risk of bias arising from measurement of outcomes (7.) Risk of bias in selection of the reported result. Signaling questions that have to be answered with yes (Y), probably yes (PY), probably no (PN), no (N) or no information (NI) help to access the risk of bias of each domain. Overall Risk of Bias was assessed using the given algorithm.

ROBINS-E is the upgraded and combined version of RoB-2.0 for randomized trials and the ROBINS-E for non-randomized trials published in 2022. It is divided into a planning phase, some preliminary questions and mainly in seven Domains. The planning phase contains defining possible confounders, the preliminary questions define the PICO values, as well as analyses for which confounder the study has been adjusted. As for the seven domains, signaling questions are asked to help identify specific concerns about potential biases. Each question must be answered with yes (Y), probably yes (PY), probably no (PN), no (N) or no information (NI). The algorithm rates Risk of bias for each of the seven domains. If there hasn't been any adjusting for confounding in the preliminary questions the algorithm already rates the study as very high risk of bias and no further questions have to be answered. Domain 1 rates high risk of bias if the results were only adjusted for age and/or sex. In domain 2 the highest quality of measuring the exposure was using a weighted eating protocol, performed over some days. This was rated as low risk, followed by questionnaires, which lead to some concerns. Domain 3 raised some concerns if participants were part of selected ethnicities or religious groups (f.e. Seventh-day Adventists) because these groups seem most likely not to be representative for western population. Domain 4 was always rated low risk of bias, because there were no post-exposure interventions. Domain 5 raised some concerns if more than 10% of the data was missing. In domain 6 low concerns were stated if the results were blood/ urine markers, as they are measured objectively in comparison to nutrient intakes arisen from questionnaires/weighted eating protocols. Concerning anthropometrics, it depends on whether they were measured (low risk) or collected in a questionnaire (some concerns). Domain 7 was usually rated low, as there were no signs for selecting results in any of the twenty studies included in this review. In the end, every domain is rated separately and the whole study is rated low risk, if all seven domains were rated low as well. Some concerns are reached, if at least one domain is rated some concerns, but none as high risk. A study is rated as high risk, if one or more domains are rated high, or a considerable amount of the domains is rated some concerns.

Table S8: Results of the certainty of evidence assesement for all conducted meta-analyses using GRADE

| Nr of studies | Nr of patients/ 95%CI | Risk of Bias | Inconsistency | Indirectness | Imprecision | Others[1] | Overall Certainty |
|---|---|---|---|---|---|---|---|
| **Protein intake (%E)** | | | | | | | |
| 3 | 175<br>-3.54 [-5.08, -2.00] | serious | serious | serious | serious | none | ⨁OOO<br>VERY LOW |
| **Carbohydrate intake (%E)** | | | | | | | |
| 3 | 273<br>5.87 [2.07, 9.67] | serious | serious | serious | serious | none | ⨁OOO<br>VERY LOW |
| **Fat Intake (%E)** | | | | | | | |
| 5 | 428<br>-1.37 [-3.46, 0.73] | serious | serious | serious | serious | none | ⨁OOO<br>VERY LOW |
| **Monounsaturated fatty acid intake (%E)** | | | | | | | |
| 2 | 253<br>-0.11 [-3.08, 2.86] | serious | serious | serious | serious | none | ⨁OOO<br>VERY LOW |
| **Polyunsaturated fatty acid intake (%E)** | | | | | | | |
| 2 | 253<br>4.21 [3.82, 4.60] | serious | not serious | serious | serious | none | ⨁OOO<br>VERY LOW |

[1] Publication Bias, Large effect, Dose response [illegible] not applicable

**Saturated fatty acid intake (%E)**

| | | | | | | | |
|---|---|---|---|---|---|---|---|
| 3 | 259<br>-6.15 [-7.30, -5.00] | serious | not serious | serious | serious | none | ⨁OOO<br>VERY LOW |

**Birth weight (kg)**

| | | | | | | | |
|---|---|---|---|---|---|---|---|
| 3 | 120<br>0.08 [-0.21, 0.38] | serious | serious | not serious | serious | none | ⨁OOO<br>VERY LOW |

**BMI (percentile)**

| | | | | | | | |
|---|---|---|---|---|---|---|---|
| 2 | 702<br>1.60 [-0.86, 4.06] | serious | not serious | not serious | serious | none | ⨁⨁OO<br>LOW |

**Fibre Intake (g/1000kcal)**

| | | | | | | | |
|---|---|---|---|---|---|---|---|
| 2 | 249<br>8.01 [6.96, 9.06] | serious | not serious | serious | serious | none | ⨁OOO<br>VERY LOW |

### Supplementary explanation for GRADE:

For each mentioned factor downgrading is possible. All studies were rated in ROBINS-E/ RoB., as far as any of the included studies in the meta-analysis is rated at least “some concerns”, there is serious risk of bias in GRADE. This was the case in all meta-analyses. Inconsistency was rated analyzing the forest plots for the directions of the confidence intervals and $I^2$ of the meta-analyses performed in RevMan5. Indirectness was serious if heterogeneous age (f.e. children mixed with adolescents) groups were analyzed. Imprecision was serious, of the population size (n) was less than 800, which was the case in all meta-analyses (46). Publication bias was not assessed, because the number of primary studies included was too small (less than 10) in all 9 performed Meta-analyses performed in this review.

Large effect and dose response were not assessed, as this would not make sense in studies concerning nutrient intake/ blood values. GRADE was only assessed for the outcomes, which are meta-analyzed in this study, because rating inconsistency would have been difficult for all the other outcomes.

https://www.embase.com/search/results?subaction=viewrecord&id=L633981015&from=export

https://www.embase.com/search/results?subaction=viewrecord&id=L22072392&from=export

(S089543561100206X), (10.1016/j.jclinepi.2011.01.0. J Clin Epidemiol [Internet]. 2021;137:265. Available from:
https://doi.org/10.1016/j.jclinepi.2021.04.014
